# Mapping Models of Tirzepatide Delivery for Obesity Management in Primary Care Settings

**DOI:** 10.64898/2026.07.24.26358784

**Authors:** Karen Coulman, Carlos Sillero-Rejon, Scott R Walter, Laura Hollands, Camilla Forbes, William Hollingworth, Jenny Lloyd, Mark Tarrant, Maria Theresa Redaniel, Andy Judge, Helen M Parretti, Richard Byng, Jonathan Pinkney

**Affiliations:** Health Economics and Health Policy Bristol, Population Health Sciences, University of Bristol, Bristol, BS8 1UD, UK; Centre for Academic Primary Care, Population Health Sciences, University of Bristol, Bristol, BS8 2PS, UK; NIHR Bristol Biomedical Research Centre, University Hospitals Bristol and Weston NHS Foundation Trust and University of Bristol, Bristol, BS1 5DS; Centre for the Psychology of Health and Wellbeing, School of Psychology, University of Plymouth, Plymouth, PL4 8AA; Faculty of Health and Life Sciences, University of Exeter, Exeter, EX1 2LU, UK; National Cancer Registry Ireland, Cork, T12 CDF7, Ireland; Musculoskeletal Research Unit, Translational Health Sciences, University of Bristol, Bristol, BS10 5NB, UK; Norwich Medical School, University of East Anglia, Norwich, NR4 7TJ, UK; Peninsula Medical School, University of Plymouth, Plymouth, PL4 8AA, UK

**Keywords:** Geographic mapping, Health policy, Health services, Primary health care

## Abstract

**Objectives:** To evaluate the early implementation of tirzepatide for the management of obesity in primary care in England, including variations in Integrated Care Board (ICB) service delivery models, access criteria, and initial prescribing activity following national policy introduction (June 2025). This paper reports the initial phase of a wider mixed methods national evaluation of tirzepatide delivery models for initial priority cohorts defined in NHS England interim commissioning guidance.

**Design:** National study combining 1) mapping of ICB implementation plans using a questionnaire, publicly available data and documents, and stakeholder discussions, and 2) analysis of English Prescribing Dataset (EPD) to investigate tirzepatide prescribing trends using interrupted time-series methods to compare monthly prescriptions before and after June 2025 (implementation start) against a synthetic control series based on semaglutide prescribing.

**Setting:** ICBs across England.

**Participants:** 23/42 ICBs (55%) responded, of which 13 also took part in stakeholder meetings. For 17 ICBs (40%), we additionally extracted information from publicly available documents.

**Main outcome measures:** Descriptions of service delivery models, access criteria, and ICB characteristics; stakeholder-reported implementation challenges; and prescribing trends over time.

**Results:** We were able to categorise models of care for 40/42 ICBs. A general practice-led delivery model was most commonly planned (18/40; 45%), followed by community/local-based delivery (8; 20%), custom models (8; 20%), and specialist weight management service (SWMS) community outreach (6; 15%). Four ICBs (10%) planned to use more than one model of care.

Most ICBs reported following national priority cohort eligibility criteria 31/39 (79%), while eight (21%) applied additional prioritisation criteria due to funding constraints. Stakeholder discussions highlighted variation in the interpretation of model definitions, variation in operationalisation, and challenges related to affordability, insufficient workforce capacity, and tight timelines, with some ICBs not yet prescribing at the time of stakeholder meetings (November 2025). Analyses of EPD showed a steady increase in tirzepatide prescribing over time, with no evidence that the June 2025 policy implementation date produced any additional increase beyond background diabetes-related trends.

**Conclusions:** Early implementation of tirzepatide prescribing in primary care has been characterised by heterogeneous service models, local adaptations of access criteria, and slow implementation of obesity-related prescribing. Findings highlight the challenges of implementing new pharmacological treatments including patient management within routine care and the need for clear guidance, realistic timelines, and system capacity. Ongoing evaluation is needed to understand how service models evolve and implications for access, equity, and outcomes.

**Summary boxes:** *What is already known on this topic:*

- Clinical trials have demonstrated the effectiveness of new obesity management medications such as tirzepatide for weight loss when combined with behavioural interventions.
- Tirzepatide has been approved for obesity management in the NHS, with a phased implementation in primary care due to concerns about affordability to expand access to treatment.
- How to deliver these treatments in routine practice, to ensure equity of access, is not yet well understood due to evolving service models and a need for real-world evaluation.

*What this study adds:*

- Early implementation of tirzepatide in primary care in England has been slow and heterogeneous, with variation in delivery models and some local adaptation of eligibility criteria in response to funding and workforce constraints.
- Between June 2025 and January 2026, there was no detectable change in prescribing attributable to obesity beyond expected background trends for diabetes, suggesting no clear early impact on obesity-related prescribing.

## INTRODUCTION

Obesity (Body Mass Index (BMI) ≥30 kg/m^2^), is a complex disease affecting nearly a third of adults in England, with serious health, societal, and economic consequences ^1–4^. Widespread prevention and treatment strategies are urgently needed. Clinical trials have demonstrated that new glucagon-like peptide-1 receptor agonists (GLP-1 RAs) such as semaglutide (Wegovy) and dual GLP-1/glucose-dependent insulinotropic polypeptide (GIP) receptor agonists such as tirzepatide (Mounjaro) achieve greater weight loss than earlier pharmacological options when combined with behavioural interventions, with average weight losses of approximately 15-21% compared with around 5-10% for previous obesity management medications ^5–7^. However, important uncertainties remain regarding adverse effects, and how these medications should be personalised, dosed, titrated and used long term in routine care. Tirzepatide, which elicits 16-18% more weight loss than placebo, was approved for overweight and obesity management by the National Institute for Health and Care Excellence (NICE) in December 2024, the first of these new medications to be approved for use outside specialist weight management services ^7–9^. Figure 1 illustrates the key evidence, policy and implementation milestones that have shaped introduction of obesity management medications in England.

**Figure 1:**
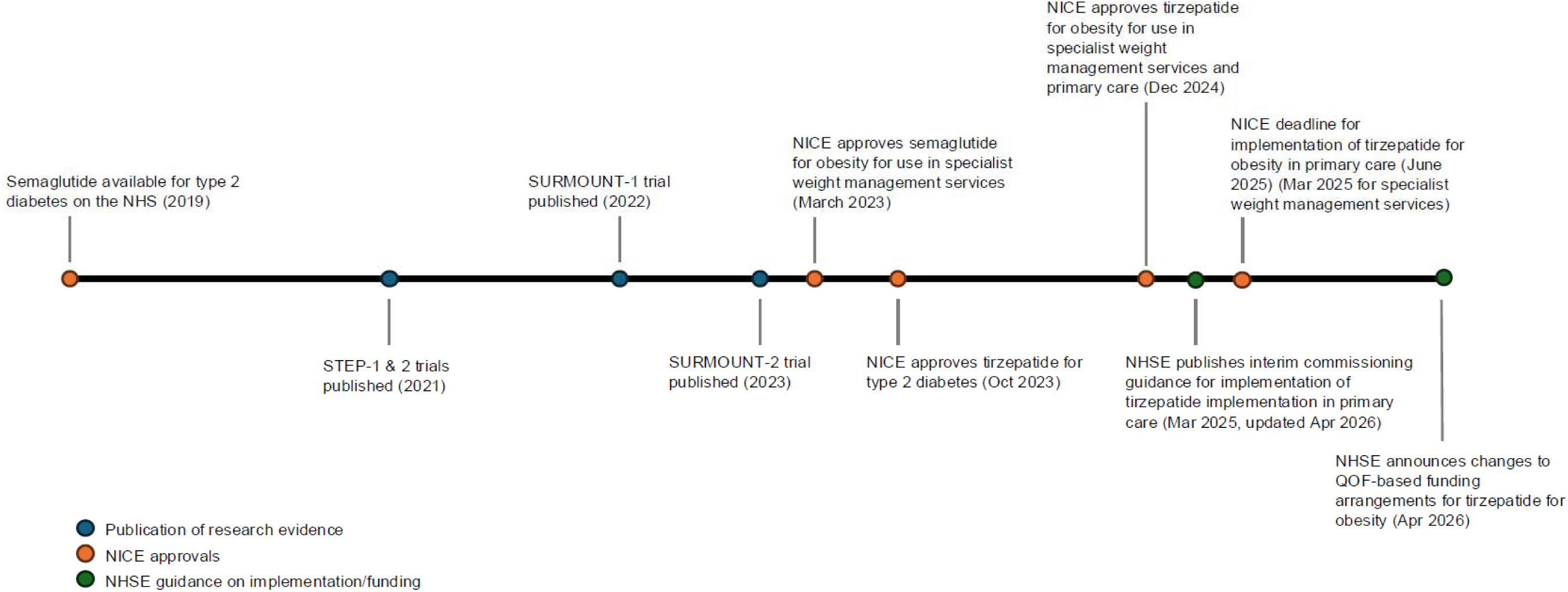
Timeline of key evidence, policy and implementation milestones for obesity management medication in England, 2019– 2026. NICE = National Institute for Health and Care Excellence, NHSE = NHS England, QOF = Quality and Outcomes Framework

It is estimated that up to 3.4 million people may be eligible for tirzepatide which has led to concerns that this would overwhelm the NHS budget if offered all at once ^8^. NHS England (NHSE) therefore submitted a variation to contract which was approved by NICE to extend the implementation period to 12 years (subsequently referred to as the ‘NICE funding variation’) ^8^. NHSE subsequently published interim commissioning guidance in April 2025 defining three priority cohorts for access to tirzepatide in primary care in the first three years of implementation (Table 1), to be provided through one of four new service models: 1) Community/local-based delivery, 2) General practitioner (GP)-led delivery, 3) Specialist weight management service (SWMS) community outreach, and 4) SWMS & GP shared care ^10^. Implementation of tirzepatide in primary care was mandated by NICE to start by June 2025 (six months from the publication of the NICE technology appraisal), with responsibility for delivery resting with Integrated Care Boards (ICBs) who allocate NHS budget and commission services for the population across geographical areas in England ^8^ ^11^.

**Table 1:** Initial priority cohorts to receive tirzepatide for obesity management in primary care as per NHS England interim commissioning guidance. ^10^

|  |  |
| --- | --- |
| Cohort 1 (months 1-12) | BMI $\geq 40 \text{ kg/m}^2$* + $\geq 4$ qualifying co-morbidities** |
| Cohort 2 (months 13-21) | BMI 35.0-39.9 $\text{kg/m}^2$* + $\geq 4$ qualifying co-morbidities** |
| Cohort 3 (months 22-36) | BMI $\geq 40 \text{ kg/m}^2$* + $\geq 3$ qualifying co-morbidities** |

BMI = Body Mass Index
\*BMI threshold reduced by $2.5 \text{ kg/m}^2$ for people from South Asian, Chinese, other Asian, Middle Eastern, Black African or African-Caribbean ethnic backgrounds
\*\*Qualifying comorbidities include: hypertension, dyslipidaemia, obstructive sleep apnoea, cardiovascular disease, type 2 diabetes

To support implementation, NHSE initially provided ICBs with funding for medication and primary care patient management costs to establish new prescribing pathways alongside centrally funded ‘wrap around’ care (i.e. behavioural support) ^10^. Such funding is not routinely provided following NICE technology appraisals. However, in April 2026 it was announced that patient management costs for tirzepatide prescribing would be incorporated into the Quality and Outcomes Framework (QOF) for 2026/27 ^10^. Under these arrangements, general practices could receive QOF payments for delivering obesity management and tirzepatide prescribing in eligible patients, representing a substantial change to the funding mechanism during the early rollout period.

England is among the first countries to integrate modern obesity management medication into a publicly funded health system outside specialist secondary care services, representing an important shift in national strategy. This approach aligns with the NHS 10-year plan’s aims of shifting care from hospital to community settings and from sickness to prevention ^12^. As an early adopter, England has the potential to generate evidence on the effectiveness and delivery of these medications at scale, informing implementation in other health systems. However, expanding access is likely to require substantial system-level changes due to the size of the eligible population and pressures on medication budgets, commissioning and primary care capacity to take up change, treatment monitoring, and behavioural support provision. Implementation has also coincided with a period of substantial financial and organisational pressures for ICBs including significant organisational change following the 2025 announcement of ICB mergers ^13^. Successful implementation will require new models of care alongside changes in clinical practices and attitudes towards obesity management. Robust evaluation of emerging service models is therefore needed to inform current and future implementation.

The National Evaluation of Weight Medication Access (NEWA) study is a national research programme designed to evaluate the emerging service models for delivering obesity management medications across primary care settings in England ^14^ ^15^. In this initial phase, we engaged with ICBs to map information on their chosen delivery models and assessed early implementation activity by investigating tirzepatide prescribing patterns.

## METHODS

### Study design

The NEWA study is a mixed-methods study evaluating the feasibility, acceptability, safety, utilisation, and clinical- and cost-effectiveness of service models for providing tirzepatide in primary care. This paper reports phase one of the NEWA study: an initial assessment of early implementation plans across ICBs and prescribing of tirzepatide in primary care since June 2025. This ‘context gathering’ phase was conducted to inform the subsequent quantitative and qualitative data collection phases of the study. This information will continue to be gathered annually during the NEWA study to gather insights on key changes as implementation progresses through each of the priority cohorts (Supplemental file 1 for study protocol). This study is reported according to the Strengthening the reporting of observational studies in epidemiology (STROBE) checklist ^16^ (Supplemental file 2).

### Data source and participants

#### Mapping information on ICBs’ implementation plans for tirzepatide

In autumn 2025, all ICBs in England were invited to complete a brief expression of interest (EoI) form (Supplemental file 1) to provide key information on their implementation plans, express interest in joining a stakeholder meeting, and in engaging with the research team on other parts of the evaluation. Information requested in the EoI included: access criteria (e.g. in line with NHSE interim commissioning guidance or different), estimated cohort volume, service model adopted (as per models outlined in NHSE interim commissioning guidance), whether they would be using the nationally commissioned behavioural support for obesity programme (BSOP) or locally provided behavioural support, and other weight management services available in the ICB area ^10^ ^17^ ^18^. Where available, we also extracted information on ICBs’ implementation plans from publicly available documents on ICBs’ websites.

#### Categorising delivery models

Delivery models adopted were categorised according to the four models of care outlined in the NHSE interim commissioning guidance ^10^. ICBs who returned an EoI form selected the model of care they had chosen to commission from the four options provided (ICB self-assessment). For ICBs who did not provide any information, we used publicly available information on ICB websites, where available, to determine the model of care (researcher assessment).

To assess the reliability of model categorisation, we compared researcher assessments of model category based on A) the free text descriptions of models of care provided in the EoI form, and B) publicly available information on ICB websites, with ICB self-assessment where available.

All researcher assessments were independently undertaken by at least two researchers (KC, CSR, SW, LH, CF) using working definitions of the four models of care provided by NHSE (Supplemental File 3), with any disagreements resolved in discussion with the wider research team.

#### Characterising Integrated Care Boards by selected model of care

To provide descriptive context on the sociodemographic, organisational and population health characteristics of ICBs adopting different models, we grouped ICBs according to their selected model and summarised a range of publicly available indicators at ICB level. These included population characteristics (registered patient population, sex distribution, and deprivation levels)^19^ ^20^, organisational characteristics (number of general practices and full-time equivalent general practitioners per 10,000 patients) ^21^, population-level indicators of potential demand for tirzepatide prescribing (prevalence of obesity and diabetes), ^22^, and tirzepatide prescribing volumes (diabetes and obesity combined) ^23^.

#### Stakeholder meetings

ICB leads who expressed interest in joining a stakeholder meeting were invited to one of two 90-minute online meetings held on Microsoft Teams. The purpose of these meetings was to introduce the NEWA team to ICBs and establish relationships, and discuss implementation plans in more detail including ICBs’ understanding of the delivery models outlined in the interim commissioning guidance, factors that influenced choice of delivery model(s), and any challenges and successes encountered in implementation. ICB leads were grouped according to model type: meeting 1 for ICBs using models 1&2 (Community/local-based delivery and GP-led delivery); meeting 2 for ICBs using models 3&4 (SWMS community outreach and SWMS & GP shared care). Key discussion points from the meetings were noted by the research team to create a descriptive summary of meeting discussions which was circulated to attendees following the meetings. Meetings were recorded and transcribed using the built-in function in Teams for cross-checking. Given the rapidly evolving landscape of obesity management medications, ICBs were invited to one of two online follow-up meetings (60-minutes) six months after the first meeting to share emerging insights from the NEWA study, discuss implementation progress, and key changes and challenges faced by ICBs.

### Patient and public involvement and engagement (PPIE)

The NEWA study has an established PPIE advisory group of eleven members with experience of living with a higher body weight and/or caring for someone living with a higher body weight. Members represent a range of socio-demographic backgrounds and regions across England. For this phase of the NEWA study, PPIE members emphasised the importance of understanding whether the public were involved in influencing ICBs’ implementation plans for tirzepatide; this was incorporated into the stakeholder discussions.

### Assessing implementation of tirzepatide using English Prescribing Dataset (EPD)

Tirzepatide (Mounjaro®) holds dual licences, allowing it to be prescribed in primary care for both type 2 diabetes and obesity. To explore early implementation, we examined national prescribing patterns for tirzepatide. We used EPD, a publicly available dataset reporting all medications prescribed by NHS primary care clinicians (e.g. GPs, nurses, pharmacists) in England and dispensed in community pharmacies ^23^. EPD includes practice-level information on the number of prescriptions issued and the total quantity prescribed. A typical prescription for tirzepatide might be for one KwikPen containing 4 doses to be taken in single weekly subcutaneous injection over a 28-day period. In the primary analysis, we used the total quantity of medicine prescribed and dispensed expressed as the number of tirzepatide injection pens. We also examined the number of prescriptions as an alternative outcome in sensitivity analyses, where one prescription may encompass multiple injection pens.

Because EPD does not record the indication for prescribing, tirzepatide prescribing data reflect use for both diabetes and obesity. We therefore included a control series to capture trends in diabetes-related prescribing, allowing us to detect any obesity-related tirzepatide prescribing over and above the control series. Semaglutide (Ozempic®) was selected as the control series because it is a GLP1-RA medication prescribed only for diabetes in primary care, serving a similar patient population and demonstrating a comparable increasing time trajectory from the start of its prescribing (January 2019), making it the most suitable available comparator for background diabetes-related prescribing trends.

### Data Analysis

#### Mapping information on ICBs’ implementation plans for tirzepatide

Quantitative information from the EoIs and publicly available datasets were analysed using descriptive statistics, summarised by delivery model categories, as specified in the NHSE interim commissioning guidance ^10^. Interrater agreement of model categorisation between ICBs and researchers was assessed using the kappa statistic (ĸ) ^24^. The level of agreement was classified as poor (ĸ ≤ 0.0), slight (ĸ 0.01– 0.20), fair (ĸ 0.21– 0.40), moderate (ĸ 0.41–0.60), substantial (ĸ 0.61–0.80), and almost perfect (ĸ 0.81–0.99) ^24^.

Free text EoI responses and stakeholder meeting notes were synthesized by LH and KC and summarised (Supplemental file 4). All ICBs invited to the stakeholder meetings received summaries via email and were asked to feedback if they did not accurately reflect the discussions.

#### Assessing implementation of tirzepatide using EPD

Monthly total quantity of tirzepatide prescribed was plotted from January 2024 to February 2026 (most recent data available prior to analysis). This range includes the NICE implementation date for tirzepatide in diabetes up to eight months after the implementation date of obesity-related tirzepatide prescribing ^25^. Trends were examined for overall prescribing as well as by dose (2.5 mg, 5 mg, 7.5 mg, 10 mg, 12.5 mg, and 15 mg). Publicly available data on population size (total patients registered with a GP practice) for England and ICBs, were used to calculate prescribing rates ^26^.

We conducted a controlled interrupted time series (CITS) analysis on monthly total quantity of tirzepatide prescribed between January 2024 and December 2025 to estimate changes before and after the policy introduction while accounting for underlying trends. The control time series included data from an equivalent period (January 2019 to December 2020) after the NICE implementation date of semaglutide for diabetes. The lack of evidence of cyclic effects or system shocks made it possible to temporally align the control series with the main tirzepatide series for analysis (Figure S1).

The control series was scaled to the tirzepatide series by minimising the sum of squares between the two series (Supplemental file 5). The policy might result in both an immediate (step change) and a more gradual (slope change) impact on tirzepatide prescribing. We report the latter based on the assumption that new service models would take time to commission and scale up.

Time was coded as a sequence of monthly intervals centred at June 2025. A binary indicator (0 = pre policy, 1 = post policy) represented the intervention periods, and another binary indicator denoted treatment (tirzepatide) versus control (time-aligned, scaled semaglutide) groups. The models included these three variables along with their two- and three way interactions. The outcomes (total quantity of pens and number of prescriptions) were modelled as a rate with population as the denominator. Given large counts and the approximately linear, rapidly increasing trends, rates were treated as continuous and modelled using linear regression, with robust standard errors. This modelling approach was applied to data for all of England combined, and separately for each ICB.

## RESULTS

### Mapping information on ICBs’ implementation plans for tirzepatide Information sources

Twenty-three of the 42 ICBs (55%) responded to the information request (20 (48%) completed the EoI form and three (7%) provided information over email) (Figure 2). The research team was able to extract some information from publicly available information on ICB websites for 17 of the 19 remaining ICBs (40%). Nineteen ICBs expressed interest in joining a stakeholder meeting, all of whom were invited to attend the stakeholder meetings. Eighteen ICB leads (representing 13 ICBs) attended one of two meetings held in November 2025. Two follow-up meetings were held in May 2026 with nine leads (representing seven ICBs) in attendance, where initial mapping findings were presented and sense-checked with attendees. Two of these seven ICBs did not attend the original November 2025 meetings but had received the November 2025 summary notes.

**Figure 2.**
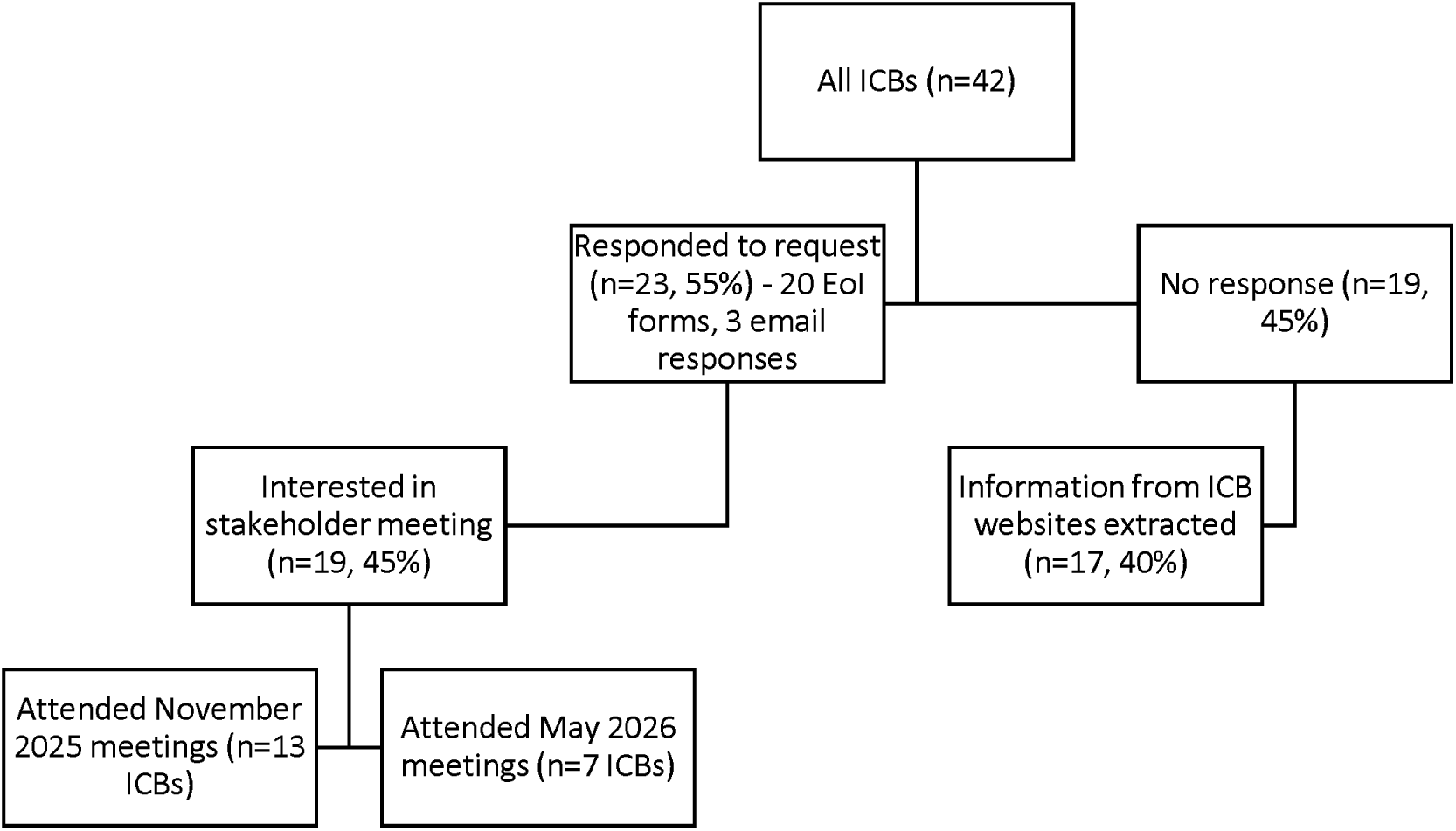
Mapping Integrated Care Boards Implementation Plans for Tirzepatide ICB: Integrated Care Board, EoI: Expression of Interest form

### Delivery models

In total we were able to categorise models for 40/42 ICBs (95%). Of these, 36 (90%) were using (or planning on using) a single model and four (10%) more than one model. The most common model was General Practice (18/40; 45%), followed by community/local-based delivery (8; 20%), custom models (8; 20%), and SWMS community outreach (6; 15%). The ‘custom’ model category included four ICBs (10%) who selected ‘custom’ model and four (10%) who selected more than one of the four NHSE-defined models of care. The SWMS & GP shared care model was not selected as a standalone model but was selected by one ICB in addition to other models of care.

ICB profiles were broadly similar across model types in terms of diabetes and obesity prevalence (Table 2). Greater variation was observed in other characteristics. For example, ICBs using custom models had the highest median estimated number of patients eligible for tirzepatide in cohort 1 (1,332; IQR 655– 1,775) and the highest mean deprivation score (22.67; SD 3.33), whereas ICBs using community/local-based delivery models had the lowest median estimated eligible population (376; IQR 330–546) and mean deprivation score (18.63; SD 4.90). ICBs using the SWMS Community Outreach model had the lowest Full Time Equivalent GPs per population (14.58, mean 3.05). The mean total quantity of tirzepatide prescribed ranged from 57.2 pens per 1,000 registered patients in ICBs with community/local-based delivery models to 81.0 pens per 1,000 in ICBs with general practice models.

**Table 2.**
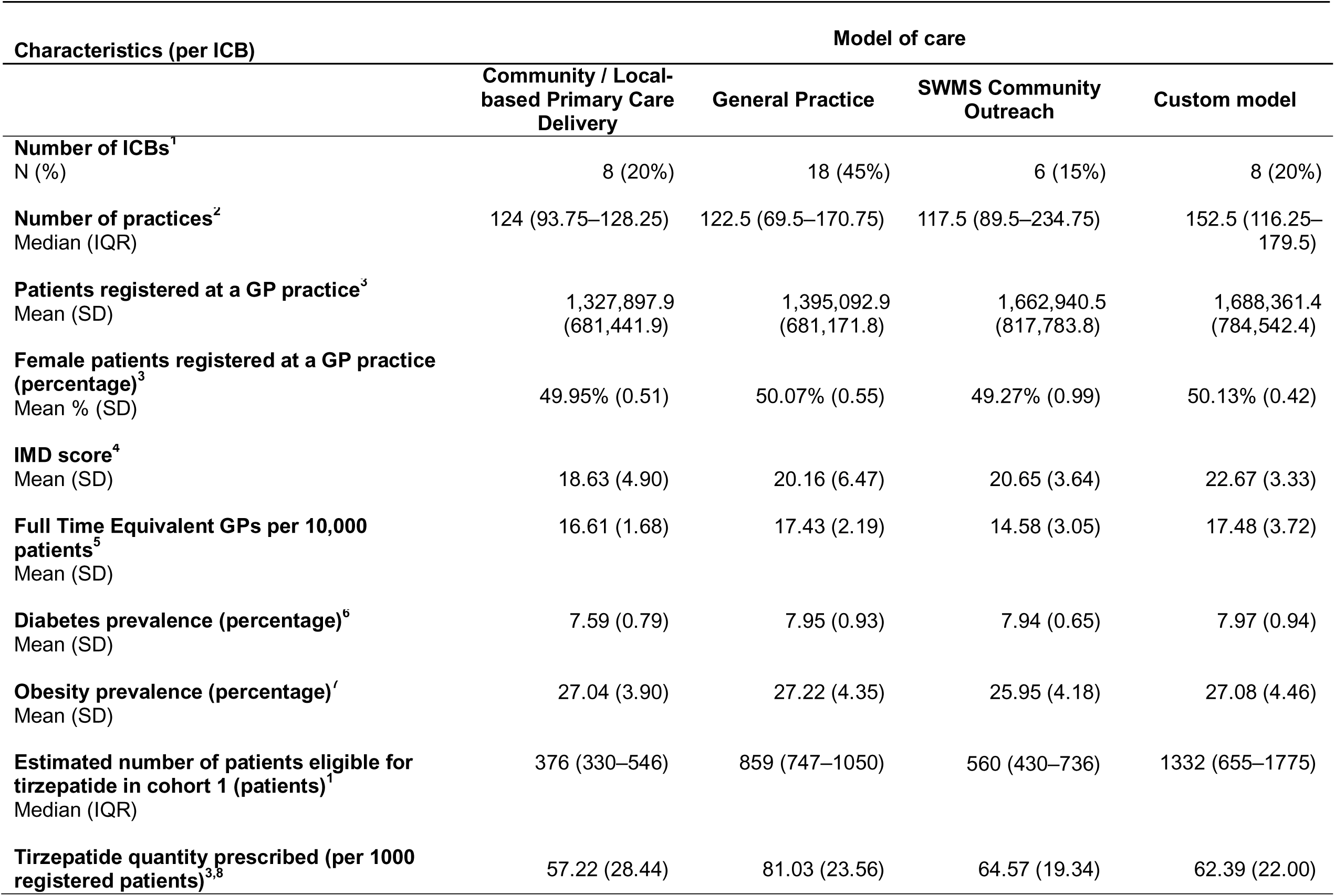
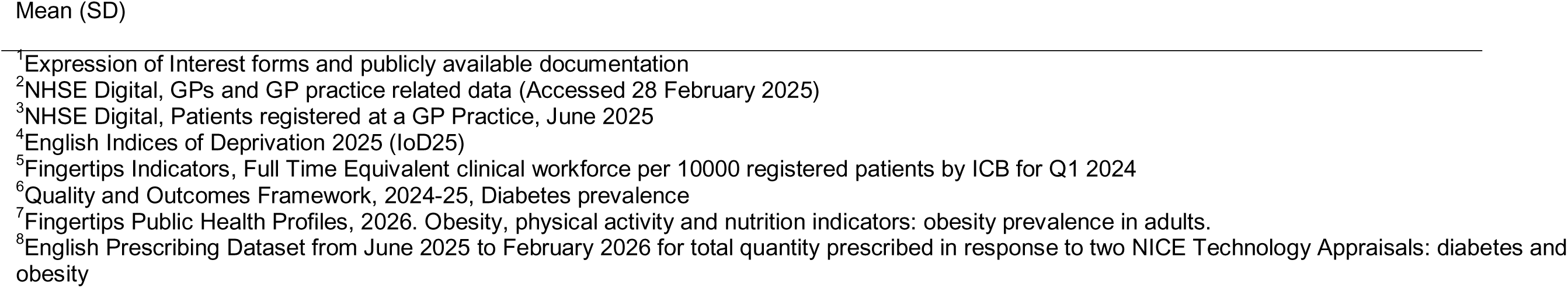
Characteristics of Integrated Care Boards grouped by model of care for tirzepatide delivery.

| Characteristics (per ICB) | Model of care |  |  |  |
| --- | --- | --- | --- | --- |
|  | Community / Local-based Primary Care Delivery | General Practice | SWMS Community Outreach | Custom model |
| **Number of ICBs<sup>1</sup>**<br>N (%) | 8 (20%) | 18 (45%) | 6 (15%) | 8 (20%) |
| **Number of practices<sup>2</sup>**<br>Median (IQR) | 124 (93.75–128.25) | 122.5 (69.5–170.75) | 117.5 (89.5–234.75) | 152.5 (116.25–179.5) |
| **Patients registered at a GP practice<sup>3</sup>**<br>Mean (SD) | 1,327,897.9<br>(681,441.9) | 1,395,092.9<br>(681,171.8) | 1,662,940.5<br>(817,783.8) | 1,688,361.4<br>(784,542.4) |
| **Female patients registered at a GP practice (percentage)<sup>3</sup>**<br>Mean % (SD) | 49.95% (0.51) | 50.07% (0.55) | 49.27% (0.99) | 50.13% (0.42) |
| **IMD score<sup>4</sup>**<br>Mean (SD) | 18.63 (4.90) | 20.16 (6.47) | 20.65 (3.64) | 22.67 (3.33) |
| **Full Time Equivalent GPs per 10,000 patients<sup>5</sup>**<br>Mean (SD) | 16.61 (1.68) | 17.43 (2.19) | 14.58 (3.05) | 17.48 (3.72) |
| **Diabetes prevalence (percentage)<sup>6</sup>**<br>Mean (SD) | 7.59 (0.79) | 7.95 (0.93) | 7.94 (0.65) | 7.97 (0.94) |
| **Obesity prevalence (percentage)<sup>7</sup>**<br>Mean (SD) | 27.04 (3.90) | 27.22 (4.35) | 25.95 (4.18) | 27.08 (4.46) |
| **Estimated number of patients eligible for tirzepatide in cohort 1 (patients)<sup>1</sup>**<br>Median (IQR) | 376 (330–546) | 859 (747–1050) | 560 (430–736) | 1332 (655–1775) |
| **Tirzepatide quantity prescribed (per 1000 registered patients)<sup>3,8</sup>** | 57.22 (28.44) | 81.03 (23.56) | 64.57 (19.34) | 62.39 (22.00) |

<sup>1</sup>Expression of Interest forms and publicly available documentation
<sup>2</sup>NHSE Digital, GPs and GP practice related data (Accessed 28 February 2025)
<sup>3</sup>NHSE Digital, Patients registered at a GP Practice, June 2025
<sup>4</sup>English Indices of Deprivation 2025 (IoD25)
<sup>5</sup>Fingertips Indicators, Full Time Equivalent clinical workforce per 10000 registered patients by ICB for Q1 2024
<sup>6</sup>Quality and Outcomes Framework, 2024-25, Diabetes prevalence
<sup>7</sup>Fingertips Public Health Profiles, 2026. Obesity, physical activity and nutrition indicators: obesity prevalence in adults.
<sup>8</sup>English Prescribing Dataset from June 2025 to February 2026 for total quantity prescribed in response to two NICE Technology Appraisals: diabetes and obesity

Figure 3 shows the ICB variation in diabetes and obesity prevalence, and the total quantity of tirzepatide prescribed between June 2025 (policy introduction) and February 2026. A marked variation in diabetes and obesity prevalence was observed across ICBs. The tirzepatide quantity prescribed ranged from ∼10 to 130 pens per 1,000, indicating strong variation in practice.

**Figure 3.**
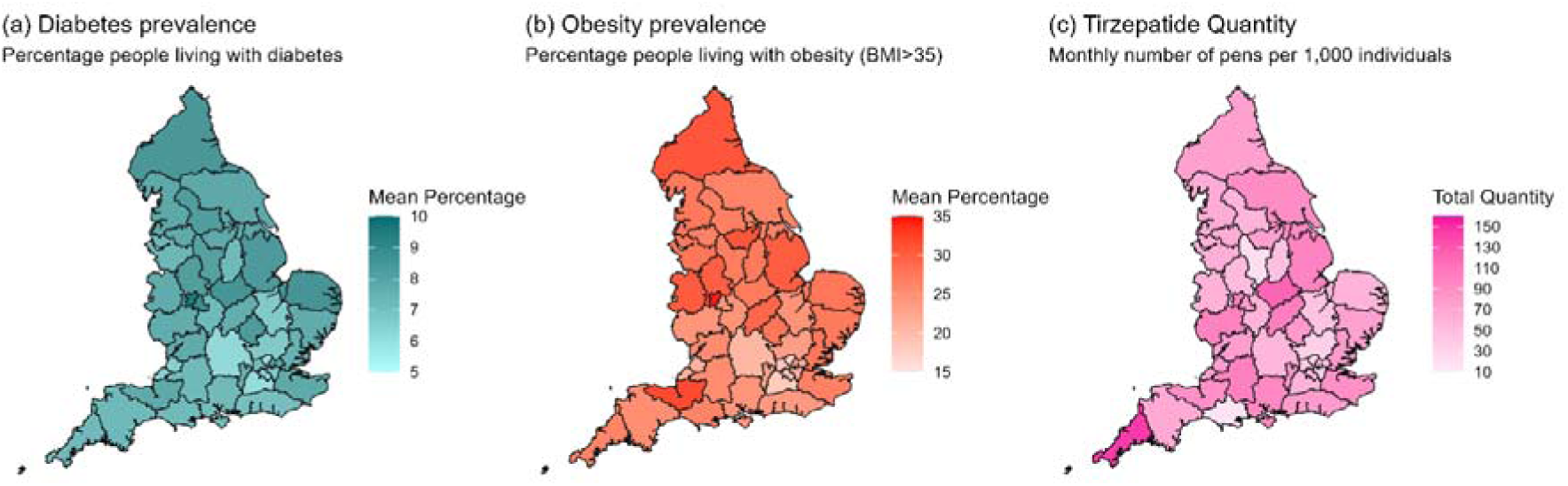
Diabetes (a) and obesity (b) prevalence and total quantity of tirzepatide prescribed from June 2025 – February 2026 (c) across Integrated Care Boards in England

At the November 2025 stakeholder meetings, ICB leads reported that model choice was shaped by several factors including affordability, existing infrastructure, equity considerations, clinical capacity, organisational changes (such as ICB mergers), and the need to establish services within tight implementation timelines. Many emphasised that their current model was not necessarily their preferred approach and expected model evolution over time, with some already planning a different model for cohort 2. Rapid timelines limited opportunities for public involvement in model design. The NICE mandated timeline was viewed as unrealistic for establishing new prescribing models with several of the ICBs in attendance reporting they were not yet prescribing.

At the May 2026 follow-up meetings, attendees discussed the recent announcement that payments for patient management of tirzepatide prescribing would be incorporated into QOF 2026/27. Attendees reported that this change to funding arrangements had differential impacts across ICBs. Some ICBs that had not used an individual general practice model for cohort 1 reported a perceived need to switch to this model for cohort 2 to align with the new funding arrangements. In some areas, the time required to procure non-General Practice-based models in cohort 1 meant that little or no prescribing had commenced before the QOF announcement, resulting in some ICBs discontinuing their originally planned delivery model(s) before, or shortly after, implementation. Some areas anticipated operating transitional or mixed models while existing patients completed treatment pathways. Views on the switch to QOF funding were mixed. Some felt it could improve access through wider primary care involvement, while others expressed concerns that variation in general practice participation could exacerbate health inequalities, particularly in more deprived areas. Participants raised concerns that the workload and reimbursement associated with QOF-funded delivery could discourage some practices from participating in tirzepatide prescribing. Others were concerned that moving from locally commissioned services to a national QOF-funded approach could reduce opportunities to specify local safety, governance and quality assurance requirements. Participants noted that areas with greater experience of tirzepatide prescribing for diabetes, and stronger supporting infrastructure within primary care (e.g. clinical pharmacy support), were in a stronger position to adapt to the QOF funding arrangements.

### Model categorisation

Researcher assessments aligned with ICB assessed models for only 14/23 (61%) or 7/23 (30%) depending on the source of information used. The level of agreement between ICB and researcher categorisations was classified as ‘slight’ (ĸ 0.10) and ‘moderate’ (ĸ 0.47) for publicly available information and text assessments, respectively.

Discussions with ICB leads highlighted variation in the interpretation of the service models in the interim commissioning guidance, with models not necessarily applied as mutually exclusive approaches. In practice, selecting a model involved choosing the label that best aligned with existing local plans. Even within models there was considerable variation in how these were implemented. For example, within the ‘General Practice delivery model’ some ICBs delivered tirzepatide via individual GP practices, while others used a small number of practices to deliver tirzepatide on behalf of a larger primary care area (e.g. GP Federation or Primary Care Network). There was also uncertainty about how such approaches should be categorised (e.g. as community/local based delivery model or General Practice).

### Access criteria, cohort size and behavioural support

Of the 39 ICBs with available information on access criteria, 31 (79%) reported following the NHSE interim commissioning guidance for cohort 1, while eight (21%) implemented locally defined criteria. These local criteria involved further prioritising specific groups within cohort 1 or putting caps on numbers that could be treated. ICB leads explained that difficulties aligning local funding allocations with the potential size of the eligible cohort 1 population were the primary reason for introducing additional prioritisation. Among the 26 ICBs that provided estimates of cohort size, the average reported cohort 1 population was 933 patients per ICB range 246– 3,000). Of 36 ICBs with information on behavioural support, all reported using nationally commissioned behavioural support with one also using local provision.

### Assessing implementation of tirzepatide using EPD

The volume of tirzepatide prescribing for diabetes across England showed a sustained and marked increase from January 2024 to June 2025 (Figures 4A & S2A). In June 2025 there were approximately 250,000 injection pens prescribed. Thereafter, with the addition of obesity prescribing, the trend continued upwards, albeit with increased variation and a notable temporary drop in August 2025. Prescribing peaked in December 2025 with over 300,000 pens prescribed. Prescribing of the lowest dose (2.5 mg), which includes initial prescribing for new patients, was broadly level after June 2025. This is consistent with a balance between the number of new patients starting tirzepatide for diabetes or obesity and the number discontinuing and/or titrating to higher doses (Figures 4B & S2B).

**Figure 4.**
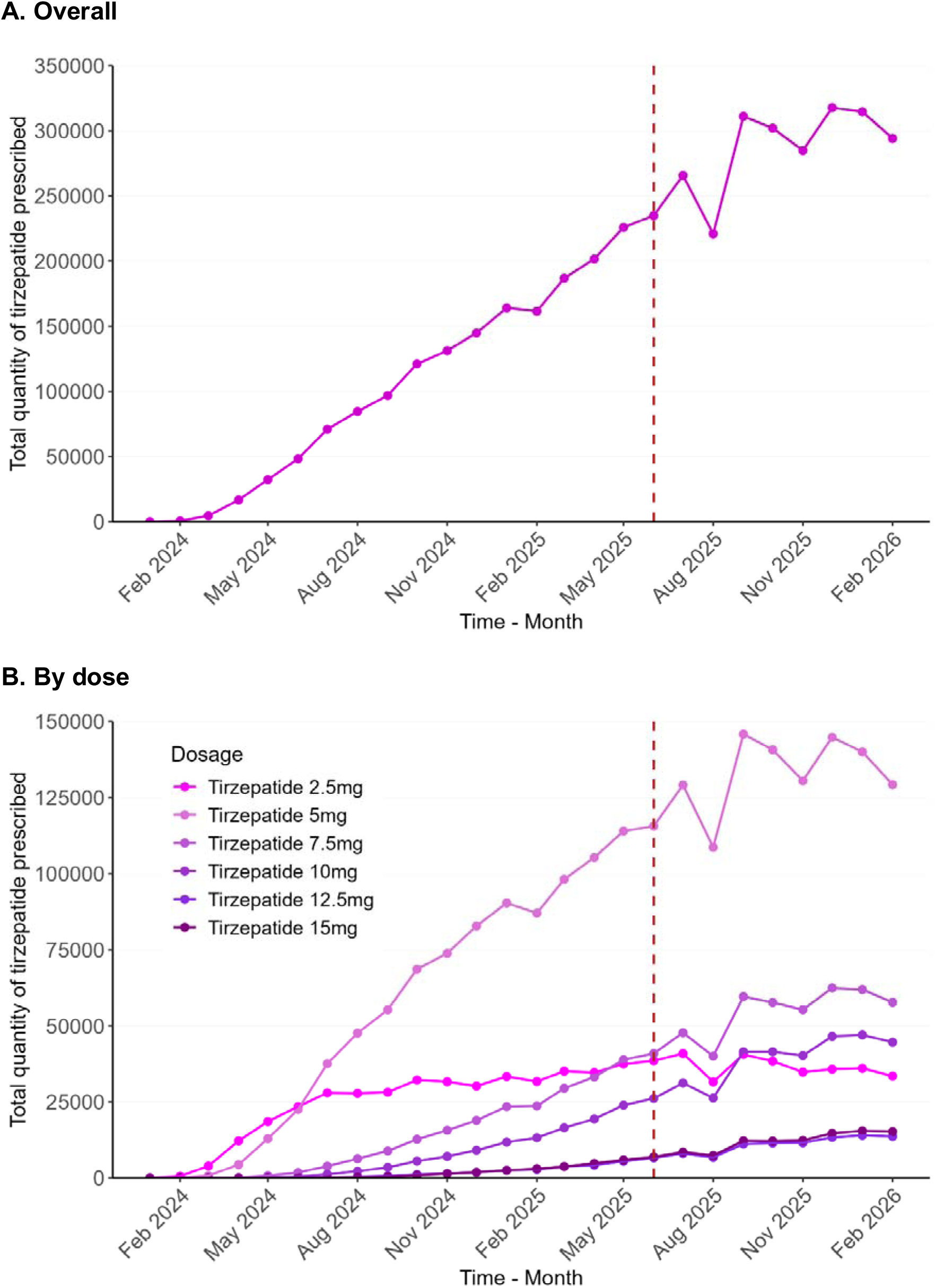
English Prescribing Data for monthly total quantity of tirzepatide prescribed - January 2024 to February 2026 The red dotted line indicates the policy introduction in June 2025 (the NICE mandated date of having access to tirzepatide for managing overweight and obesity in place).

The CITS analyses did not show any clear evidence of an increase in total quantity of tirzepatide prescribed above background diabetes-related rates after policy introduction. This was true both nationally (Figure 5, Table S1) and across individual ICBs (Figure 6). Among the few statistically significant ICB-level slope change estimates, all were negative indicating post-policy tirzepatide prescribing rates below expected based on background diabetes-related trends (Figures 6 & S4). Sensitivity analysis of the number of prescriptions also did not show evidence of a post-policy increase in tirzepatide prescribing (Table S1, Figure S3).

**Figure 5.**
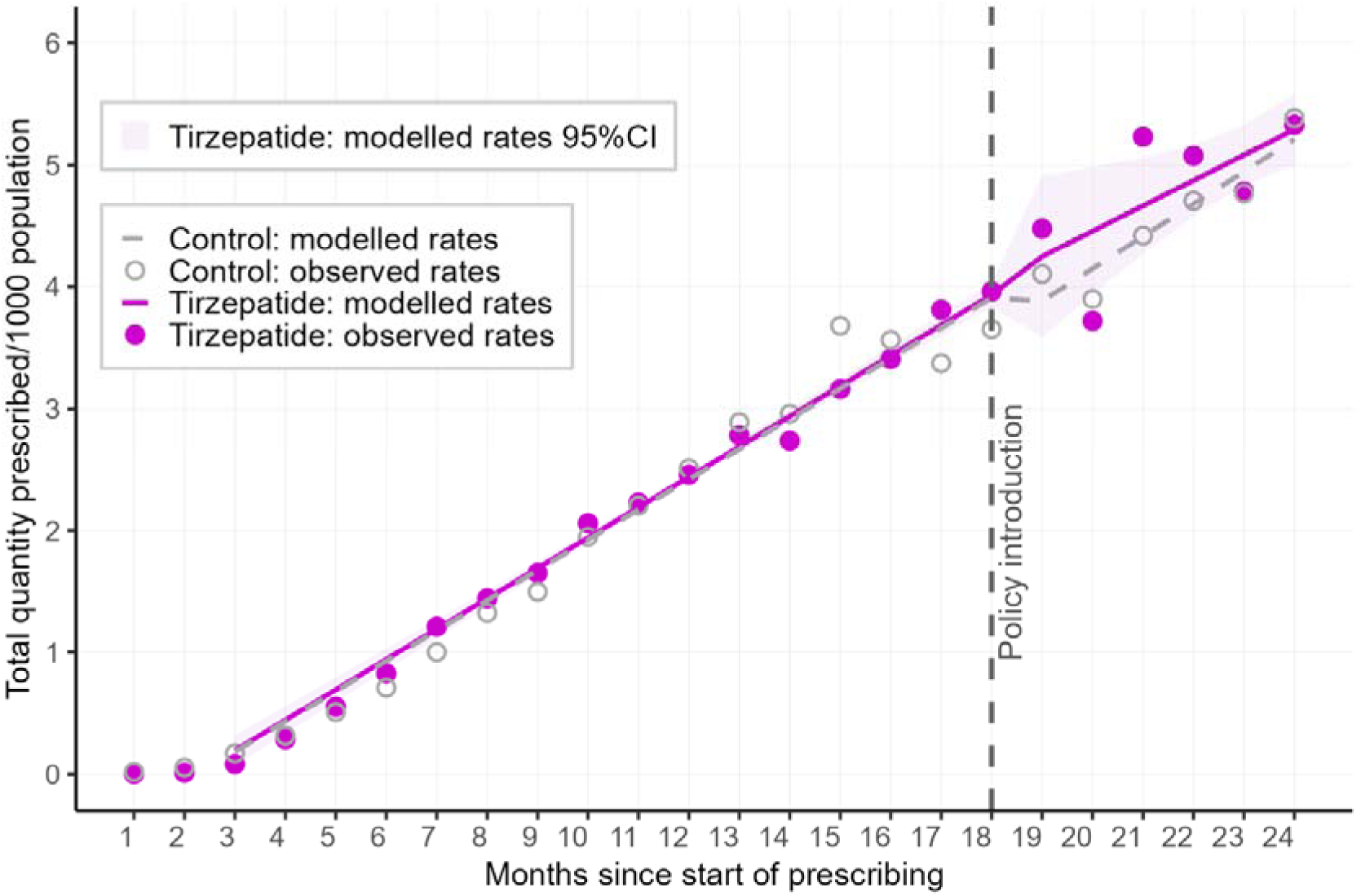
Controlled interrupted time series results for the rate of tirzepatide prescribing (total quantity prescribed per 1000 population) compared to a control series (time-aligned, scaled semaglutide rates), based on English Prescribing Data for all of England combined. Note: Start of prescribing for tirzepatide in primary care was January 2024, and for semaglutide, January 2019.

**Figure 6.**
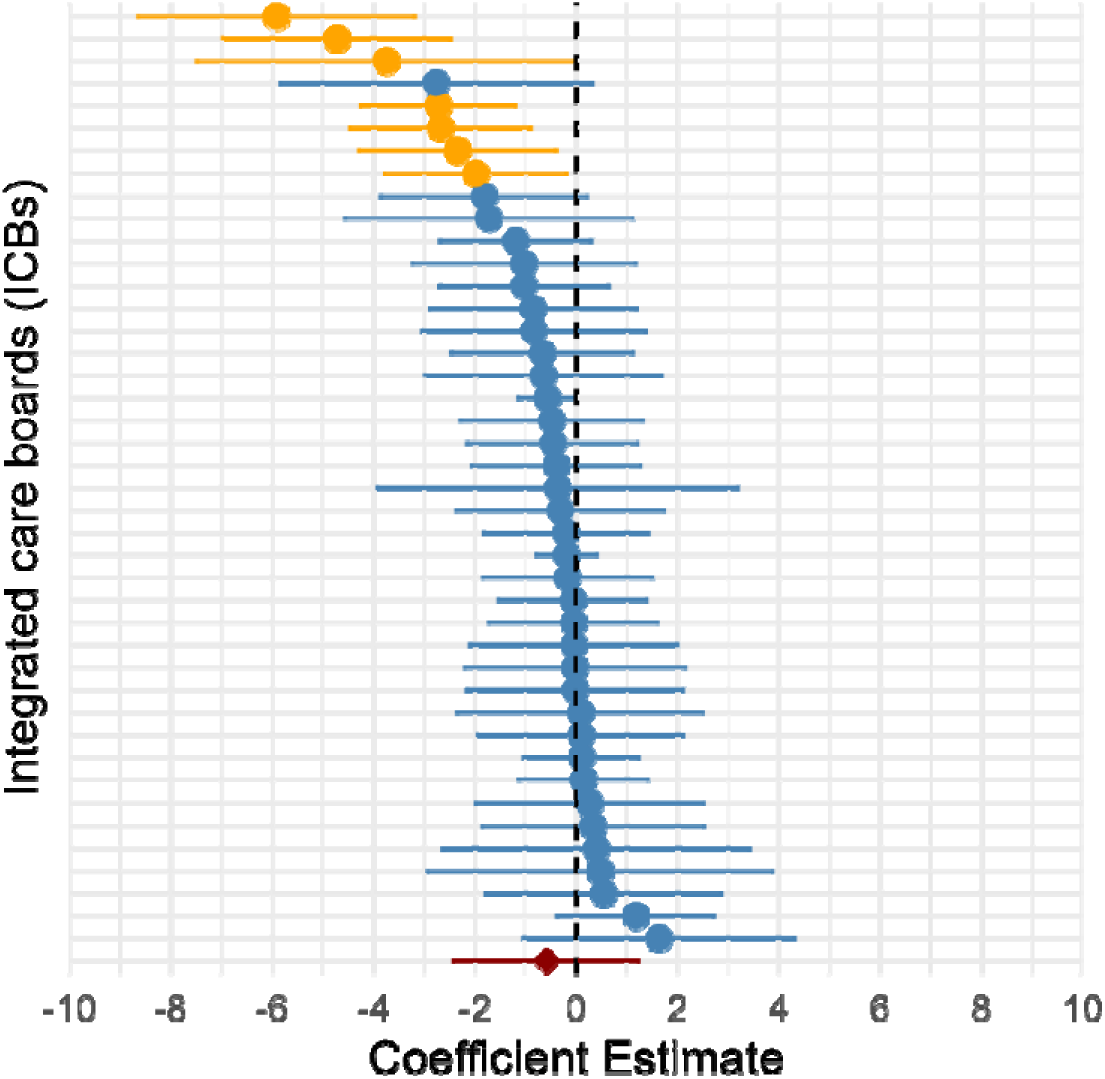
Controlled Interrupted Time Series results assessing longer-term slope change of the total quantity of tirzepatide prescribed over and above background trends of diabetes-related prescribing, by ICB. Each point and 95% CI line represents an Integrated Care Board. Values below zero indicate a slower increase in tirzepatide prescribing rates relative to the control series, while values above zero indicate faster relative increase. Yellow lines and dots indicate a 95% confidence interval that does not overlap the null value (zero), where there is evidence of a difference in trend in quantity prescribed relative to the control series.

## DISCUSSION

We identified substantial heterogeneity in the models used to deliver tirzepatide prescribing. There was variable local interpretation of the four approaches to service models outlined by NHS England. Prescribing activity was slow to initiate, as reflected in both self-reported accounts and in analyses of routine prescribing data. Stakeholders attributed this to short timelines for establishing new service models, commissioning and implementation challenges, and difficulties responding to evolving policy and funding arrangements. We also found evidence that local population and organisational characteristics may have influenced model selection, with custom models more commonly adopted in areas with larger eligible populations and higher levels of deprivation. Together, these findings suggest that early implementation has been shaped by local system pressures, with implications for equity and consistency of access.

A key strength of this study is that it represents the first phase of a national three-year evaluation of tirzepatide implementation in primary care, providing an early, system-level overview to inform subsequent phases. The combination of questionnaires, stakeholder meetings, and national prescribing data enabled triangulation and a more comprehensive understanding of implementation across England. However, several limitations should be considered. Findings are based on early, self-reported implementation experiences from a subset of ICBs and publicly available sources, which may be incomplete or inconsistently reported. Prescribing analyses relied on aggregated datasets that do not capture individual-level clinical outcomes or adherence. Furthermore, obesity-related prescribing volumes were small relative to prescribing for diabetes, which may have limited our ability to detect changes above background prescribing trends. The prescribing analysis also relied on a control derived from diabetes-related semaglutide prescribing to enable estimation of obesity-related tirzepatide prescribing activity beyond expected background diabetes-related trends. As with any quasi experimental design, there are inherent limitations due to differences in launch timing, prescribing environments, and eligibility criteria. Nevertheless, the inclusion of the control strengthens the causal estimation of the policy implementation effect relative to analysing tirzepatide prescribing without a control.

The findings reflect an early stage of implementation, and both service models and prescribing trends are likely to have evolved since data collection, as indicated by our latest stakeholder meeting discussions in May 2026. Policy developments, including the planned incorporation of tirzepatide prescribing into the QOF for the 2026/27 GP contract, with the aim of supporting GP practices to consistently identify adults living with obesity and promote equitable access to obesity management interventions including tirzepatide, may further influence implementation and choice of delivery models ^27^ ^28^. Mapping of implementation plans, and prescribing trends will be repeated annually during the NEWA study to capture changes over time.

To our knowledge, NEWA is the first study to evaluate the early implementation of tirzepatide in primary care. Existing literature is largely limited to policy guidance, commentaries and news reports on implementation challenges. Our findings are consistent with emerging reports, including in *The BMJ,* highlighting challenges in the early rollout of tirzepatide in primary care, such as constrained commissioning arrangements, uncertainty around eligible population sizes, and incomplete national rollout into early 2026 ^28^ ^29^. Concerns have also been raised that the phased rollout and prioritisation approach may exacerbate existing health inequalities and contribute to a two-tier system of access to obesity treatment ^30^. Our findings extend these reports by identifying how tight implementation timelines, limited clinical and commissioning capacity, organisational change within the NHS, and evolving national policy, including changes to funding and commissioning arrangements, have shaped local delivery models and their evolution over time. These factors may help explain the slow rollout and low levels of obesity-related prescribing observed during the early implementation period.

Several factors may explain the patterns of prescribing observed. Although NICE technology appraisals do not typically include additional funding for implementation, the introduction of tirzepatide for obesity was accompanied by dedicated funding from NHS England, creating both opportunities and complexity for local decision-making. The need to rapidly establish new prescribing pathways across primary care, SWMS, and behavioural support provision may have exceeded what was feasible within the initial timelines, particularly given the substantial organisational changes affecting ICBs during this period. Previous qualitative work has also highlighted concerns among GPs regarding capacity, resources, and the practicalities of integrating GLP-1 receptor agonists into routine primary care ^31^. The absence of a clear increase in obesity-related tirzepatide prescribing in our analysis compared to the diabetes prescribing trajectory is likely to reflect a slow start to prescribing for obesity management, supported by our stakeholder meeting insights. We also observed a temporary decline in prescribing in August 2025, immediately before a substantial increase in the private UK price of tirzepatide from September 2025. Although this pricing change related to the private market, it may have influenced prescribing patterns through wider effects on demand, supply or movement between private and NHS prescribing. These changes may have affected prescribing patterns around the intervention period, complicating interpretation of the time-series analysis.

Our descriptive analyses also suggest that implementation decisions may have been influenced by local context. While obesity and diabetes prevalence were broadly similar across model types, ICBs adopting custom models tended to have larger estimated eligible populations and higher levels of deprivation than those using community/local-based models. This may reflect adaptation of delivery models to local population needs, capacity, or service infrastructure. However, given the observational nature of the data and the early stage of implementation, no conclusions can be drawn regarding the relative effectiveness of different delivery approaches.

These findings have important implications for commissioners and policymakers. Clearer guidance on service models, realistic implementation timelines, and support for system coordination is likely to be critical to enable consistent and equitable rollout across the country. Without this, there is a risk of continued variation in access between ICBs. Although incorporation of tirzepatide into QOF aims to support consistent identification and equitable access to treatment, as it is a voluntary scheme there is a concern, highlighted through stakeholder insights, that its impact may vary across areas and could risk reinforcing existing inequalities, particularly where participation or implementation differs between general practices ^10^.

Further research is needed to assess how different service models influence access, outcomes, and clinical- and cost-effectiveness as implementation progresses. Ongoing data collection within the NEWA study, including repeated mapping of implementation plans and prescribing trends, will be important to capture how services evolve over time and in the context ICB mergers in 2026 and 2027 ^13^. There is also a need to examine the impact of local prioritisation decisions on equity and to identify delivery approaches that are scalable and sustainable within routine care.

Drawing on insights from this initial phase, we have developed three working definitions of cohort 1 delivery models (*individual general practice*, *general practice ‘at scale’*, and *separate weight management services*). These reflect how services were enacted in practice, rather than the models as described in interim commissioning guidance and will be refined as further data are collected and prescribing increases across the country, to ensure they continue to capture how models of care evolve in real-world settings.

## Ethics statement

Phase one of the NEWA study received approval from the Faculty of Health Sciences Research Ethics Committee at the University of Bristol (ref 25475). ICB stakeholders were engaged as experts to provide contextual information on the introduction of tirzepatide prescribing in primary care across England, informing subsequent work packages. The study used organisational-level data, publicly available information including aggregated national prescribing data.

## Data sharing statement

Data are publicly available or fully reported. Analysis code is available at: https://osf.io/n7vau/overview

## Acknowledgements

We thank the members of the NEWA PPIE group for their ongoing advice and support on the NEWA study. We are grateful to the Integrated Care Board colleagues who shared information on local implementation plans for tirzepatide, attended stakeholder meetings, and generously gave their time to support this evaluation. We particularly acknowledge their contribution during a period of significant organisational change and service development within the NHS.

## Author contributions

KDC co-led the conception, design and funding acquisition for the NEWA study, contributed to data collection, analysis, and interpretation of findings and led the drafting of the manuscript.

CSR contributed to the conception and design of the study, data collection and analysis including prescribing data analyses, interpretation of findings, and drafting and revision of the manuscript.

SRW contributed to the conception and design of the study, data collection and analysis including prescribing data analyses, interpretation of findings, and reviewing and revising the manuscript.

LH contributed to data collection, analysis, interpretation of findings, and reviewing and revising the manuscript.

CF contributed to data collection, analysis, interpretation of findings, and reviewing and revising the manuscript.

JL contributed to the conception, design and funding acquisition for the NEWA study, data collection, analysis, interpretation of findings, and reviewing and revising the manuscript.

WH contributed to the conception, design and funding acquisition for the NEWA study, data analysis, interpretation of findings, and reviewing and revising the manuscript.

MT contributed to the conception, design and funding acquisition for the NEWA study, data collection, analysis, interpretation of findings, and reviewing and revising the manuscript.

MTR contributed to the conception, design and funding acquisition for the NEWA study, data analysis, interpretation of findings, and reviewing and revising the manuscript.

AJ contributed to the conception, design and funding acquisition for the NEWA study, data analysis, interpretation of findings, and reviewing and revising the manuscript.

HMP contributed to study design, interpretation of findings, and reviewing and revising the manuscript.

RB contributed to the conception, design and funding acquisition for the NEWA study, interpretation of findings, and reviewing and revising the manuscript.

JP co-led the conception, design and funding acquisition for the NEWA study, contributed to interpretation of findings and reviewing and revising the manuscript.

All authors critically reviewed the manuscript for important intellectual content, approved the final version for publication, and agree to be accountable for all aspects of the work.

KDC is the guarantor.

## Transparency statement

Dr Karen Coulman (the manuscript’s guarantor) affirms that the manuscript is an honest, accurate, and transparent account of the study being reported; that no important aspects of the study have been omitted; and that any discrepancies from the study as planned (and, if relevant, registered) have been explained.

