## Supplemental file 1 for "Mapping Models of Tirzepatide Delivery for Obesity Management in Primary Care Settings"

### National Evaluation of Weight-medication Access: Mapping the Implementation of Tirzepatide for the Management of Obesity in the National Health Service

#### NEWA Mapping

**Protocol version number: 2**

**Date: 10 December 2025**

**Funder:** National Institute of Health and Care Research (NIHR162959)
**Sponsor: University of Bristol**
**Sponsor reference:**
**FREC reference: 25475**
**IRAS reference: NA**

**Protocol version number and date:**

| Version | Date | Change |
|---|---|---|
| 1.0 | | N/A |
| 2.0 | 10th December 2025 | Addition of interrupted time series analysis of publicly available English Prescribing Data |

NEWA
NATIONAL EVALUATION OF WEIGHT-MEDICATION ACCESS

University of BRISTOL

**Principal Investigators: Karen Coulman and** **Jonathan Pinkney**

**Co-applicants:**

| | | |
|---|---|---|
| **Andrew Judge** | Professor of Translational Statistics | University of Bristol |
| **Carlos Sillero Rejon** | Senior Research Associate | University of Bristol |
| **Carly Hughes** | Honorary consultant (GP) | The Fakenham Medical Practice |
| **Helen Parretti** | Consultant Clinical Associate Professor | University of East Anglia |
| **Jenny Lloyd** | Senior Lecturer in Public Health Research | University of Exeter |
| **Jessica Munafo** | Clinical Psychologist | Southmead Hospital |
| **Jonathan Pinkney** | Professor of Endocrinology, Honorary Consultant in Weight Management | University of Plymouth |
| **Karen Coulman** | Senior Research Fellow and Weight Management Dietitian | University of Bristol |
| **Katalin Bagi** | Research Portfolio Manager | NHS BNSSG ICB |
| **Kenneth Clare** | PPI/E Lead | PPI Representative |
| **Louise Lacey** | PPIE | PPI Representative |
| **M. Theresa Redaniel** | Head of Research and Analysis | National Cancer Registry Ireland |
| **Mark Tarrant** | Professor of Psychology | University of Plymouth |
| **Marsha Johnson** | Costing Specialist | University of Bristol |
| **Nysha Givans** | Doctoral Student | PPI Representative |
| **Richard Byng** | Professor in Primary Care Research | University of Plymouth |
| **Scott Walter** | Research Fellow | University of Bristol |
| **Will Hollingworth** | Professor of Health Economics | University of Bristol |

**Study Statisticians:** Scott Walter and Andrew Judge

**Study co-ordinator:** Karen Coulman

#### KEYWORDS

Obesity, Weight Management, Tirzepatide, GLP1

#### GLOSSARY OF ABBREVIATIONS

| | |
|---|---|
| **BMI** | Body mass index |
| **SWMS** | Specialist weight management service |
| **ICB** | Integrated Care Board |
| **NHSE** | NHS England |
| **WM** | Weight management |

### INTRODUCTION

#### BACKGROUND

Glucagon-like peptide 1 receptor agonists (GLP1-RA) are a new family of anti-obesity medications, including Semaglutide and Tirzepatide. Of these medications, Tirzepatide (Mounjaro©) has been shown to elicit more weight loss than placebo and Semaglutide (Wegovy©) in a clinical trial (1) and more than Semaglutide in an observational setting (2). These medications offer more effective obesity treatment when combined with behavioural interventions (1,3,4) and require wraparound care. Recognising the demand and need for wider access, NICE guidance for Tirzepatide has opened prescribing, in conjunction with behavioural support, to primary care (5). NHS England (NHSE) is thus implementing four alternative service models for accessing Tirzepatide, necessitating a comprehensive evaluation to guide future rollout.

##### MODELS OF CARE

Based on the NHSE interim commissioning guidance (5), approximately 220,000 patients in three priority cohorts will be treated over the initial three-year implementation period (June 2025 – May 2028). Priority cohort 1: BMI ≥40 kg/m$^2$ and ≥4 'qualifying' co-morbidities (n≈30,000, months 1-12) cohort 2: BMI 35.0-39.9 kg/m$^2$ and ≥4 'qualifying' co-morbidities (n≈40,000, months 13-21), cohort 3: BMI ≥40 and 3 'qualifying' co-morbidities kg/m$^2$ (n≈150,000, months 22-36) – see Appendix B. In all cases, the BMI threshold is reduced by 2.5 kg/m$^2$ for people from South Asian, Chinese, other Asian, Middle Eastern, Black African or African-Caribbean ethnic backgrounds. Patients will be treated within one of four implementation models:

1. Community / Local-based delivery model
2. General Practice delivery model
3. Specialist weight management services provision of a community outreach delivery model

4. Specialist weight management services, Community & General Practice shared-care model.

Each Integrated Care Board (ICB) will choose the model(s) they wish to adopt. Within each model, ICBs can choose to provide wraparound care through local provision or through a central funded NHSE provision.

A mixed methods NIHR funded research evaluation of the new models of care will be undertaken by the research team. Establishing key stakeholder contacts and gathering important contextual information on the nature of the NHS rollout is first needed to inform the subsequent research components of the evaluation.

#### PROJECT AIM

This project aims to establish key stakeholder contacts, gather contextual information from stakeholders, and map information related to the ICBs' chosen model(s) of care in the implementation of weight medication access. We will also aim to synthesise publicly available information related to weight medication. Insights will inform subsequent research components of the wider evaluation.

#### OBJECTIVES

For each priority cohort, we aim to:

- Establish key contacts at each ICB and NHSE.
- Understand the model(s) of care to implement anti-obesity medication access in each ICB.
- Describe the different models of care for the delivery of anti-obesity medication.
- Describe the wraparound care provided alongside the models of care in each ICB.
- Describe the implementation dates and delivery dates in each ICB.
- Describe the models of care against ICBs characteristics, including anticipated patient numbers, patient characteristics, other weight management services in the area, etc.
- Assess the extent of tirzepatide prescribing in England

#### METHODS

##### STUDY DESIGN

Descriptive service evaluation, interrupted time series study

##### STUDY SETTING

Integrated Care Boards, NHS England, England.

#### STUDY DURATION

Three years, June 2025 – February 2028. It is expected that the mapping exercise will be done three times alongside the expected cohorts: cohort 1 (June-September 2025), cohort 2 (June-September 2026) and cohort 3 (February-May 2027). Therefore, each mapping exercise will last approximately three months.

#### INFORMATION COLLECTION

First, from NHSE, we will identify key contacts and publicly available policy documents. Similarly, we will do this at each ICB. Each ICB will also be asked to complete an Expression of Interest form (see Appendix A) providing details of their chosen service model. ICB representatives will also be invited to meetings to improve our understanding of the key contextual factors underpinning the NHS rollout to inform how we refine the research design for subsequent work packages. The meetings will occur at mutually agreed timepoints, likely at six-monthly intervals as suggested by ICB representatives. The information gathered from this process will encompass:

- Priority cohorts to be treated with Tirzepatide (and if different to interim commissioning guidance).
- Service model adopted and key features of the model(s).
- Wraparound care characteristics and provider.
- Anticipated numbers of patients within priority cohorts to be treated.
- Other locally available WM services within each ICB (e.g. whether a T3SWMS is available).
- Intended implementation dates for each priority cohort.

From secondary, publicly available sources we will obtain:

- ICBs' geographical boundaries from the Office for National Statistics Open Geography portal (6)
- GP practices and locations from NHS England Digital and House of Commons Library (7,8)
- Population-level demographics and obesity prevalence obtained from the national GP profiles (e.g. age, sex, deprivation, ethnicity) (6)
- Anonymised, open-source prescribing data from NHS England: The English Prescribing Dataset (9).

The secondary data will be obtained annually.

#### DATA MANAGEMENT

Data will be stored on password-protected servers located at the University of Bristol and appropriately backed up. Only approved study investigators will have access to data storage folders.

#### DATA ANALYSIS

Descriptive statistics such as counts and proportions will be represented at ICB-level for all relevant variables via tabulation and graphical outputs, including geospatial maps (11). This will be done for characteristics of models of care and WM services (model chosen, inclusion of wraparound care, source of wraparound care, availability of T3SWMS), demographic information (e.g., deprivation, age, gender), obesity prevalence, and GP practices.

Analyses of prescribing data will use an interrupted time series framework analyse monthly counts of prescriptions for tirzepatide (Mounjaro) before and after the implementation of primary care prescribing of tirzepatide for obesity (June 2025 = the intervention). Firstly, we will model the change in the distribution of tirzepatide doses using up to 18 months of data prior to the intervention (assumed to be diabetes-related prescriptions only in pre-intervention period) and at least six months after the intervention (a mixture of diabetes- and obesity-related prescriptions).

We will also model the overall monthly count of tirzepatide prescriptions (for all doses combined) and compare changes following introduction of obesity-related prescribing to a control series. Controls will be monthly counts of semaglutide (Ozempic), a similar GLP-1 receptor agonist to tirzepatide prescribed for diabetes in primary care. Given the rapid increase in prescribing counts following the introduction of each drug, the two series will be aligned as per the date they were approved for primary care prescribing. Changes in the main time series (Monjaro counts) over and above any changes in the control series will be considered as attributable to the intervention.

Although the policy for primary care prescribing of tirzepatide commenced in June 2025, in practice the start dates varied across ICBs. Where ICB-specific prescribing start dates are known, will carry out controlled interrupted time series analysis at ICB level, then combine those results using meta-analysis methods.

### ETHICS, REGULATORY ISSUES AND DISSEMINATION

#### RESEARCH ETHICS

Approval has been provided by Faculty of Health Sciences Research Ethics Committee

#### CONFIDENTIALITY

All data will be handled according to the principles of the Data Protection Act, the GDPR, and the University of Bristol's Research Governance and Integrity Policy (November 2019).

Data will be either publicly available or will be at the level of ICB and relate to characteristics of service provision. There will be no individual-level data or personal identifiers.

#### STUDY SPONSORSHIP

Sponsorship of this research project is being sought from the University of Bristol.

#### CONFLICT OF INTEREST

There are no specific conflicts of interest in relation to the research team. This project will follow the NIHR Conflict of Interest Policy for Funding and Awards.

#### PATIENT AND PUBLIC INVOLVEMENT AND ENGAGEMENT (PPIE)

The wider project will establish a pool of 10 PPIE contributors, prioritising groups with higher obesity rates and who may be less likely to access weight management services. The PPIE group will be involved in design, analysis, and contributing to research summaries and creative dissemination outputs. This protocol and related documents have been reviewed by the projects PPIE co-leads.

#### DISSEMINATION

This information will be rapidly summarised in a brief report that will be shared with NHSE as soon as complete. This will be updated annually during the lifetime of the project and will provide essential information that will feed into other work packages in the full evaluation. We will seek publication in open-access journals and interactive tools that would be publicly available for ICBs, NHSE, patients, the general public and other stakeholders, following principles of open research. The timescale of report production and updating is proposed as follows:

| Milestone | Dates |
|---|---|
| Cohort 1 analysis | June-September 2025 |
| Cohort 2 analysis | June-September 2026 |
| Cohort 3 analysis | February-May 2027 |

NEWA
NATIONAL EVALUATION OF WEIGHT-MEDICATION ACCESS

University of BRISTOL

NEWA
NATIONAL EVALUATION OF WEIGHT-MEDICATION ACCESS

University of BRISTOL

### APPENDIX

#### APPENDIX A. EXPRESSION OF INTEREST FORM

List of involved institutions

| | | |
|---|---|---|
| University of Bristol | Leeds Beckett University | University of Exeter |
| University of Plymouth | University of East Anglia | Bristol, North Somerset and South Gloucestershire Integrated Care Board |

[Date]

### National Evaluation of Weight-medication Access (NEWA)

#### Expression of Interest

Dear ICB representative

The National Evaluation of Weight medication Access (NEWA) is commissioned and funded by the National Institute for Health and Care Research (NIHR). It is being led by researchers at the Universities of Bristol, Plymouth, Exeter and Leeds Beckett, and hosted by Bristol, North Somerset and South Gloucestershire Integrated Care Board (ICB). The project will assess the feasibility, acceptability, safety, effectiveness, and cost-effectiveness of new service models for providing Tirzepatide. The evaluation focuses on initial priority cohorts and its findings will inform future implementation of service models to promote equitable access to anti-obesity medication.

A fundamental aspect underpinning the design of the evaluation is to have a clear understanding of the models of care to be used by each ICB. The purpose of this form is

- to establish a key contact at your ICB,
- to gather initial information about the model of care in your ICB, and
- to gain an indication as to the extent of the ICBs involvement as a stakeholder in this evaluation.

Thank you so much,

Yours sincerely,

Dr Karen Coulman (University of Bristol) & Prof Jonathan Pinkney (University of Plymouth)

#### ICB Contact

| | | |
|---|---|---|
| **ICB NAME**<br>*Please indicate the name of your ICB.* | Click or tap here to enter text. | |
| **ICB Contact**<br>*Please nominate a person from your ICB for NEWA to contact in the future regarding this study.* | Name | Click or tap here to enter text. |
| | Role | Click or tap here to enter text. |
| | Email | Click or tap here to enter text. |
| | Phone number | Click or tap here to enter text. |

#### ICB Characteristics

| | |
|---|---|
| **FIRST COHORT CHARACTERISTICS**<br>*Please, describe the eligibility criteria for the first cohort to access tirzepatide (Mounjaro®)* | ☐ As per the NHS England interim commissioning guidance (BMI 40+ and at least 4 'qualifying' co-morbidities) |
| | ☐ Other (please describe): Click or tap here to enter text. |
| **FIRST COHORT VOLUME**<br>*Estimated total number of eligible patients in the first cohort* . | Click or tap here to enter text. |
| **ACCESS DATE(S) FOR SUBSEQUENT COHORTS**<br>*Please tell us what date(s) you anticipate access will be widened to other cohorts, if known at this stage.* | Click or tap here to enter text. |
| **MODEL OF DELIVERY**<br>*Use the checkboxes to indicate which model(s) the ICB will be implementing. If choosing "other", please describe it.* | ☐ **Model i**: Community / Local-based Primary Care Delivery |
| | ☐ **Model ii**: General Practice |
| | ☐ **Model iii**: Specialist Weight Management Service (SWMS) Community Outreach |
| | ☐ **Model iv**: SWMS & General Practice Shared Care |
| | ☐ **Other**: Custom model |
| **MODEL DESCRIPTION**<br>*In a few words, please describe your chosen model(s) of care in more detail, including the reasoning behind the choice. If there are any local documents that describe your model of care and you are able to share these, please include these in your email response.* | Click or tap here to enter text. |
| **WRAPAROUND CARE**<br>*How will your ICB implement wraparound care alongside tirzepatide (Mounjaro®)? Use the checkboxes.* | ☐ We will use the nationally commissioned (NHS England)wraparound care. |
| | ☐ We will use our own wraparound care |
| **WRAPAROUND DESCRIPTION** | Click or tap here to enter text. |

NEWA
NATIONAL EVALUATION OF WEIGHT-MEDICATION ACCESS

University of BRISTOL

| | |
|---|---|
| *If using the nationally commissioned wraparound care, please state the provider you will be using in your area.*<br><br>*If using your own local wraparound care, what will this consist of? How will it be offered to patients? How will it be implemented?* | |
| **OTHER WEIGHT MANAGEMENT SERVICES**<br>*Please tell us briefly what other weight management services are available in your area (e.g. Tier 3/specialist weight management service). If this information is publicly available, you may wish to include a link here.* | Click or tap here to enter text. |

#### Further contact between ICB and evaluation team

Having a clear understanding of the models of care implemented by each ICB is an important part of the evaluation. In some case we may need to get in touch to gather additional information or clarify responses provided in this form. Please indicate if you are happy for us to contact you for further information as needed.

☐ I am happy for the evaluation team to contact the ICB (via the details provided above) if necessary for further information to fulfil the NEWA evaluation.

An online stakeholder meeting will be held as part of our stakeholder engagement process. The focus of this meeting will be on gathering shared understandings of the different service models being used in the rollout, including how these models were originally conceptualised and designed, and important contextual information on a national and local level. It is expected to take up to two hours and involve a range of discussion activities, e.g. breakout groups. Please indicate your willingness to participate in this meeting below.

☐ I am willing to participate in an online stakeholder engagement meeting.

The evaluation aims to work closely with four ICBs as case study sites. This would involve in-depth qualitative research (interviews, focus groups) with people living with obesity and with health professionals. A key feature of our proposal is a 'researcher in residence' approach at our case study ICB sites, together with community researchers to support the recruitment of underrepresented groups to the research. We would seek NHS honorary contracts for our researchers to facilitate this approach, enabling our researchers to be fully embedded within case study sites and build essential relationships with clinical teams, commissioners, and local communities. Community researchers will be local to each case study site area and will also be part of our study-specific Patient and Public Involvement group. Please indicate if your ICB would potentially be interested in being a case study site below. **Please note that this is just to gauge interest and does not commit you to anything at this stage.** Should you indicate interest below, the research team will then provide further details on what is involved and would set up a time to discuss this with you. Case study sites will be selected to ensure different service models and population socio-demographics are included.

☐ The ICB is interested in having a discussion about potentially being a case study site for the evaluation.

| | | | |
|---|---|---|---|
| **Name** | Click or tap here to enter text. | | |
| **Signed** | ☐ Checking this box will be accepted in place of a signature if you are submitting this form via email | **Date** | Click or tap to enter a date. |
| PEASE EMAIL THIS FORM BACK TO **** | | | |

NEWA
NATIONAL EVALUATION OF WEIGHT-MEDICATION ACCESS

University of BRISTOL

DATA PROTECTION: Please note that personal information recorded in this form will be treated with an appropriate level of confidentiality and handled in accordance with GDPR and the Data Protection Act 2018. This information will be made available only to those within the study team who need access in order to fulfil the evaluation. Information on service models will be summarised at ICB level and fed into published reports.

#### APPENDIX B

Eligibility criteria to receive Tirzepatide treatment as per NICE guidelines:

| Cohort | Eligible from | BMI | Comorbidities |
|---|---|---|---|
| I | June 2025 | ≥40* | At least 4 out of: Hypertension, dyslipidemia, obstructive sleep apnoea, cardiovascular disease, type II diabetes. |
| II | June 2026 | ≥35* & <40* | At least 4 from the above list |
| III | February 2027 | ≥40* | At least 3 from the above list |

*A lower BMI threshold to be used (usually reduced by 2.5 kg/m2) for people from South Asian, Chinese, other Asian, Middle Eastern, Black African or African-Caribbean ethnic backgrounds
