## Supplemental file 3 for "Mapping Models of Tirzepatide Delivery for Obesity Management in Primary Care Settings"

NHSE summary of the models:

- **General Practice model**

In this model, the whole pathway is situated within the GP setting. This includes the initial assessment, follow-up appointments, and continued titration and monitoring. The patient is referred to behavioural wraparound care (BSOP) by a referrer in the General Practice - this could be the GP, a nurse, other HCP, or admin staff.

- **GP and SWMS Shared Care model**

The GP reviews whether the patient is eligible for tirzepatide, then refers onwards to the SWMS Tier 3 service for the initial service assessment, who assume care for the initiation and drug titration for the initial phase of treatment. Once the patient has reached the maximum tolerated dose and is safe to transfer to GP prescribing, a discharge summary and transfer of prescribing is actioned. Ongoing titration and monitoring is then led within the GP. The patient is referred to behavioural wraparound care (BSOP) by a referrer in the SWMS Tier 3 service.

- **SWMS Community Outreach model**

The GP reviews whether the patient is eligible for tirzepatide, then refers onwards to the SWMS community outreach service for the initial service assessment. The clinician-led follow-up appointment also occurs in this setting, as well as ongoing titration and monitoring. The patient is referred to behavioural wraparound care (BSOP) by a referrer in the SWMS outreach service.

- **Community/local-based Delivery model**

The GP reviews whether the patient is eligible for tirzepatide, then refers onwards to the community/local-place based service for the initial service assessment. The clinician-led follow-up appointment also occurs in the community service, as well as ongoing titration and monitoring. The patient is referred to behavioural wraparound care (BSOP) by a referrer in the community service.
