## Supplemental file 4 for "Mapping Models of Tirzepatide Delivery for Obesity Management in Primary Care Settings"

**Summary of key points from NEWA November 2025 stakeholder meetings:**

Thirteen ICBs joined the two stakeholder meetings held in late November.

Most reported that local models for tirzepatide prescribing were shaped by what could realistically be mobilised within tight timescales, rather than reflecting the long‑term ideal pathway. Many expect their models to evolve over time. Decisions were influenced by existing T3 capacity, local partnerships, and anticipated future configurations, including clustering with neighbouring ICBs.

GP‑led delivery appealed in areas where GPs already manage obesity‑related comorbidities, though concerns were raised about training, managing demand, and low patient numbers per practice. Some areas addressed this by clustering delivery through PCNs or GP Federations.

Variations between central funding allocations and locally identified eligible numbers were highlighted, with some areas sub-prioritising within cohort 1 to manage this, and some highlighting concerns around equity. Some felt that eligibility criteria were not aligned with local clinical expectations.

Timelines were widely perceived as challenging, with many areas only recently commencing delivery or yet to begin. The importance of maintaining a strong focus on prevention and behavioural support within the broader obesity pathway was emphasized.

Topics you hope will be addressed through the evaluation:

- Addressing health inequality
- The impact on co-morbidities, not just weight
- How to shape delivery models in the future
- Return on investment

**Summary of key points from NEWA May 2026 stakeholder meetings:**

Seven ICBs joined the two stakeholder meetings held in May 2026.

Stakeholders discussed the transition from locally commissioned cohort 1 models to QOF-supported primary care delivery, and the differential impact this had across ICBs. Some ICBs that had not used a general practice-led model for cohort 1 reported switching to this model for cohort 2 to align with QOF. The time required to procure new (non-General Practice-based) delivery models in cohort 1 meant some ICBs had not been able to deliver any prescribing (or very little) before the QOF announcement was made with some subsequently closing their cohort 1 models before or soon after the start of their implementation. Some areas anticipated operating transitional or mixed models while cohort 1 patients complete treatment pathways.

Views on the impact of QOF were mixed. Some felt it could improve access to tirzepatide through wider primary care involvement, while others raised concerns around GP perceptions that QOF may not represent fair payment for work, as well as a lack of patient safety pathways that could be written into locally commissioned services.

Attendees discussed the potential impact on health inequalities, including concerns that variation in practice participation could result in unequal access to treatment, particularly in deprived and underserved communities. There was also discussion about variation in prescribing confidence across primary care, with some areas describing training and education initiatives to support implementation.

Comments were made that areas that were less concerned about workload implications seemed to be those with a strong track record of tirzepatide prescribing for diabetes with good infrastructure for additional roles in general practice, e.g. clinical pharmacists.

Attendees also highlighted uncertainty around the transition between cohorts and the challenge of meeting demand where funding had not been sufficient to cover all eligible patients.

Stakeholders mentioned that the national behavioural support had not had as much uptake as expected and there were challenges with access to face-to-face provision in one area in attendance. Discussions highlighted uncertainty about the implications for patients who decline, discontinue or are unable to access behavioural support, and a need to better understand engagement with behavioural support alongside tirzepatide treatment.
