## Supplemental file 5 for "Mapping Models of Tirzepatide Delivery for Obesity Management in Primary Care Settings"

**Derivation of synthetic controls based on semaglutide prescribing rates**

The inability to distinguish tirzepatide prescribing for diabetes versus obesity in the English Prescribing Dataset (EPD) and the recent implementation of tirzepatide prescribing in primary care suggests the need for a control series that acts as a counterfactual for recently rolled out diabetes-only tirzepatide prescribing. The nearest approximation of this is semaglutide prescribing since this is also a GLP1-RA medication prescribed for diabetes in primary care and where the initial implementation period is captured within the EPD.

The rollout of semaglutide prescribing began in January 2019 and hence does not temporally align with the Tirzepatide rollout from January 2024 (Figure S1A). However, the monthly time series for tirzepatide and semaglutide both start essentially from zero and follow an approximately linearly increasing trajectory over the first two years following implementation. There was no evidence of cyclic (e.g. seasonal) effects or any major shifts due to background factors in either series. We use these features to justify the temporal alignment of the two series, assuming that the evolution of the semaglutide series would have followed the same linearly increasing trajectory starting in 2024 as it did in 2019 (Figure S1B).

Tirzepatide and semaglutide rates were expected to follow similar adoption processes given the similarities of the drugs, the prescribing context and the rollout. Given the nearly linear pre-intervention trends over time for both drugs starting essentially from zero, albeit with differing gradients, a multiplicative scaling factor estimated from the pre-intervention period was used to scale the control trajectory to mimic the main (tirzepatide) series as closely as possible (Figure S1C). This assumed the two series to be proportional and have structurally similar growth patterns.

For a log-linear approach, it would have been possible to use the time-aligned semaglutide series as a control by applying a standard CITS parameterisation, where the slope difference between the tirzepatide and control series would have been a constant on the log scale. However, the rapid linear growth and large counts of the series meant that such models fit the data poorly. Linear regression was used instead as this provided a more appropriate fit to the prescribing rates.

Our approach considered a single control series and aimed to find an optimal multiplicative weight applied to the values of that series to minimise its distance from the main series based on data from the pre-intervention period. This is distinct from a typical synthetic control approach which would use a weighted combination of multiple potential control series.

Consider the sum of squares between the main time series and a weighted control series:

$$S\left( w \right)=\sum_{i=1}^{n} (y_{i}-wx_{i})^{2}$$

where the $y_{i}$ and $x_{i}$ are the monthly values of the main (tirzepatide) and control (time-aligned semaglutide) series’, respectively, over the pre-intervention period $\left( i=1,\ldots,n \right)$, and $w$ is a scalar weight. We aim to find the value of $w$ that minimises $S(w)$ using a least-squares approach.

We differentiate $S(w)$ with respect to $w$

$$\frac{dS}{dw}=\sum_{i=1}^{n} 2(y_{i}-wx_{i})(-x_{i})=-2\sum_{i=1}^{n} x_{i}(y_{i}-wx_{i}).$$

Setting the derivative to zero

$$-2\sum_{i=1}^{n} x_{i}(y_{i}-wx_{i})=0$$

$$\sum_{i=1}^{n} x_{i}y_{i}-w\sum_{i=1}^{n} x_{i}^{2}=0.$$

Solving for $w$ then gives the following estimator

$$\hat{w}=\frac{\sum_{i=1}^{n} x_{i}y_{i}}{\sum_{i=1}^{n} x_{i}^{2}}$$

The control series values are then given by $x_{i}^{*}=\hat{w}x_{i}$ over the full analysis period, $i=1,\ldots,n,\ldots,N$.

A simple linear regression of $y_{i}=\beta_{0}+\beta_{1}x_{i}^{*}+\epsilon_{i}$ for the pre-intervention period gave an $R^{2}$ of 0.98 for both the total quantity prescribed per 1,000 population and for the number of prescriptions per 1,000 population, indicating that the assumption that the two series are proportional is well supported by the data.
