## Supplemental Table S1 and Figures S1-S4 for "Mapping Models of Tirzepatide Delivery for Obesity Management in Primary Care Settings"

**Figure S1**. Illustration of control series construction - rates of total quantity prescribed per 1,000 population for semaglutide and tirzepatide from the English Prescribing Dataset.

| **A)** Rates on original time scale | |
| --- | --- |
| 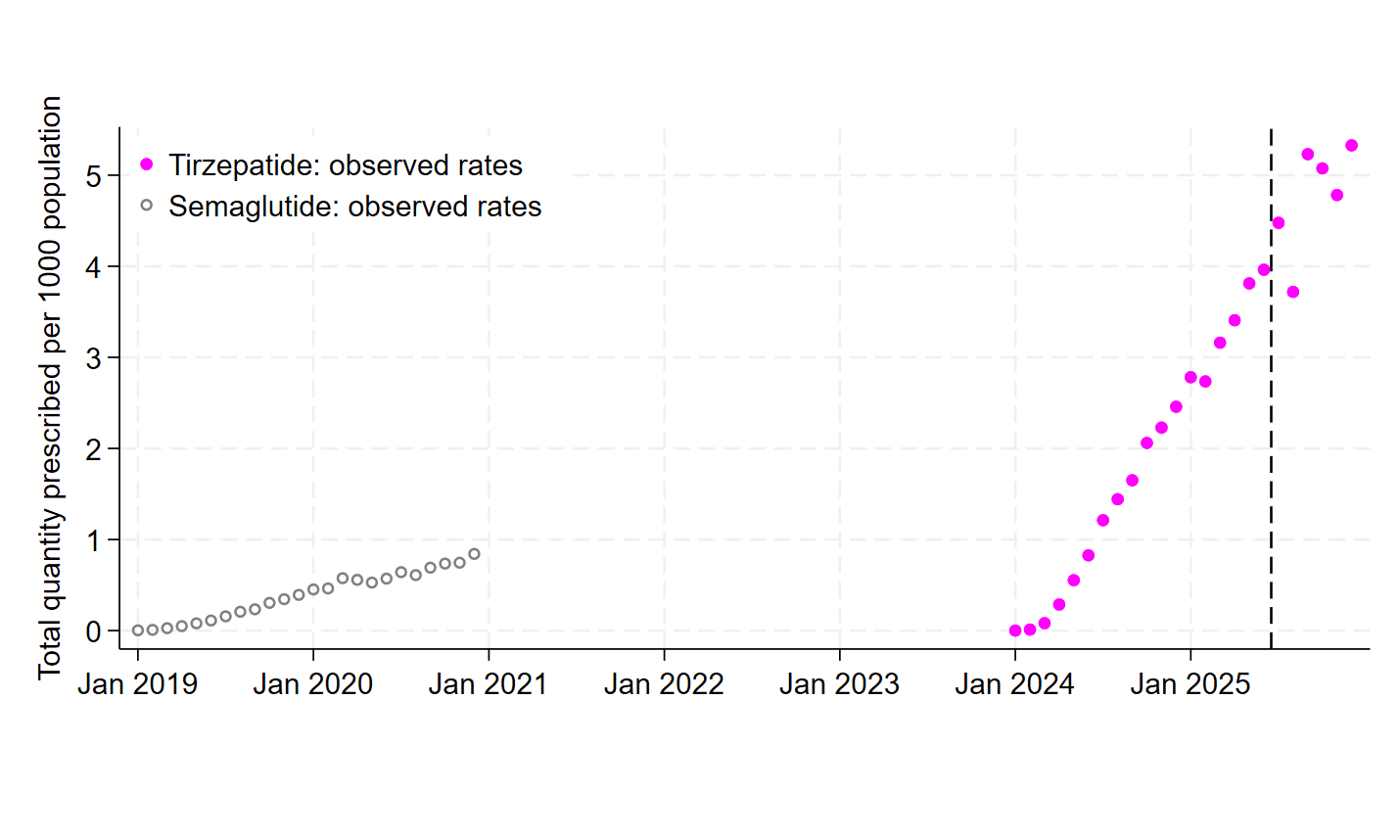 | |
| **B)** Time-aligned rates | **C)** Time-aligned and scaled rates |
| 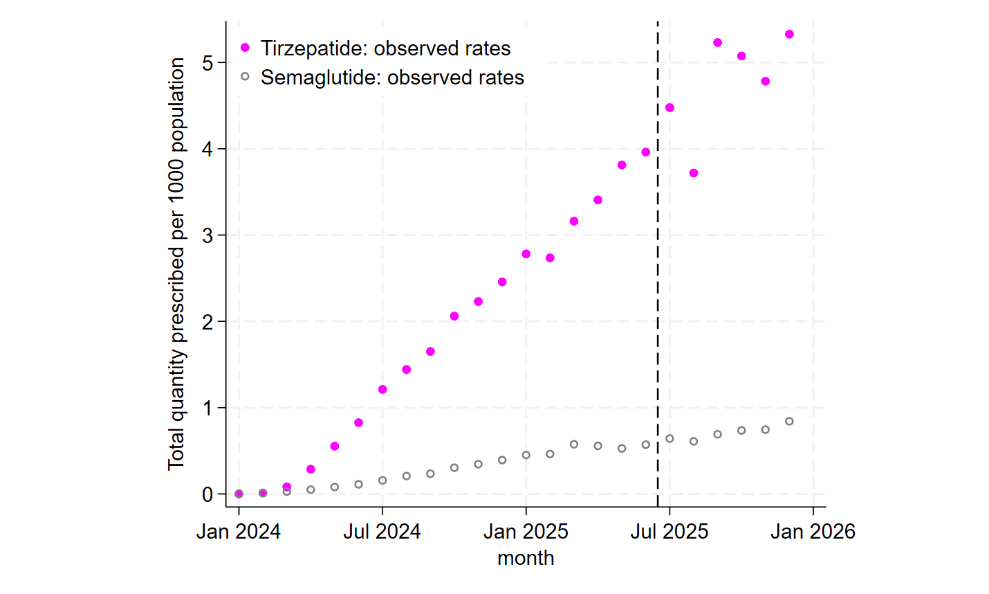 | 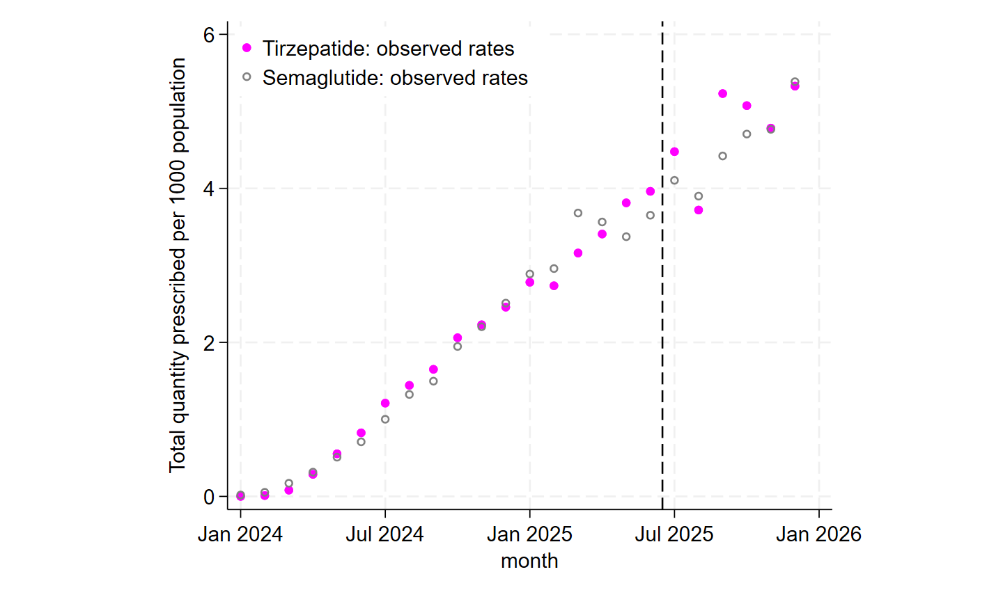 |

**Figure S2.** English Prescribing Data for monthly number of tirzepatide prescriptions – January 2024 to February 2026

**A. Overall**

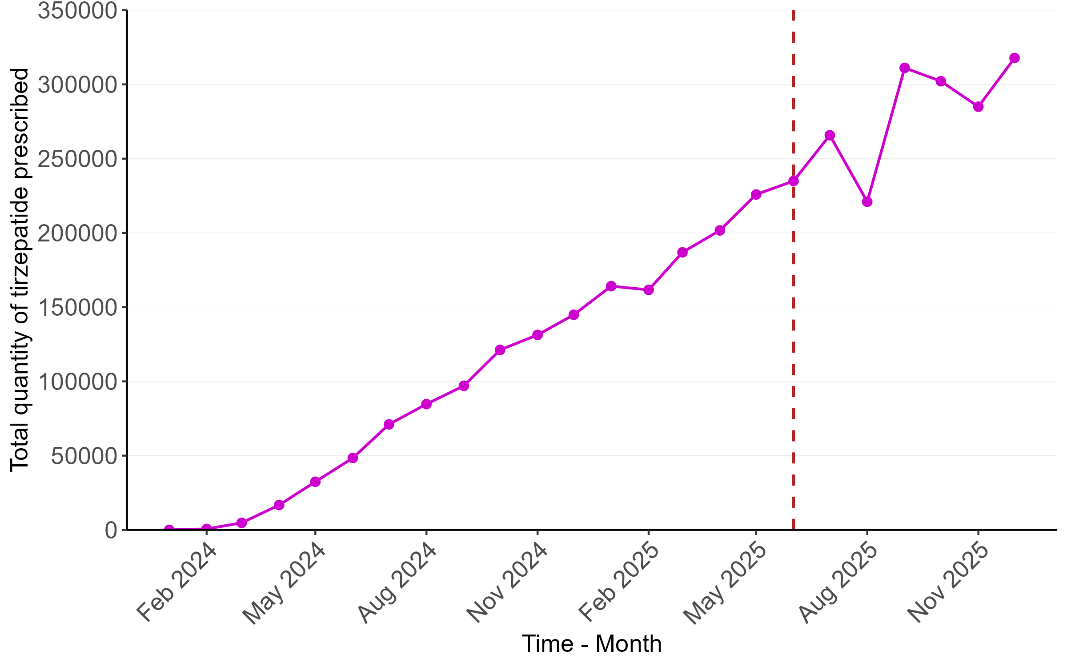

**B. By dose**

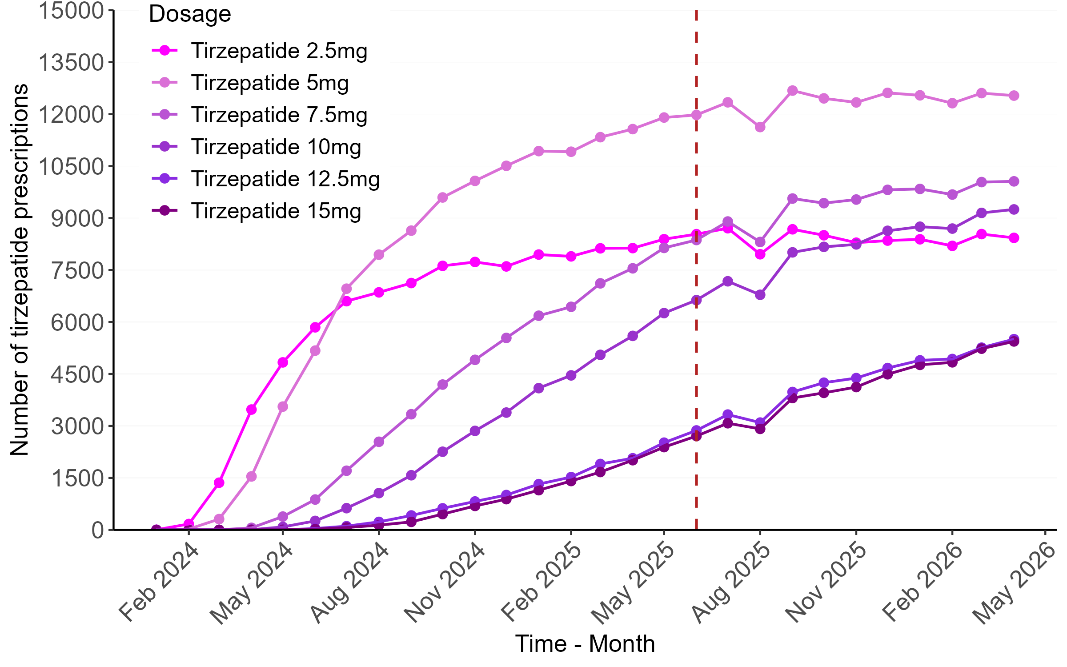

**Table S1** Controlled interrupted time series estimates of the impact of introducing the obesity medication pathway on prescription rates for tirzepatide, using semaglutide as a control and based on the English Prescribing Dataset. Estimates derived from linear regression of prescribing rate per 1000 population.

| **Parameter** | **Estimate** | **95% CI** | **P-value** |
| --- | --- | --- | --- |
| **Total quantity prescribed per 1000 population** |  |  |  |
| Intervention (post vs. pre) | -0.283 | -0.710, 0.145 | 0.20 |
| Drug (tirzepatide vs. semaglutide) | 0.025 | -0.244, 0.295 | 0.85 |
| Time | 0.248 | 0.226, 0.271 | <0.001 |
| Time x intervention | 0.017 | -0.081, 0.115 | 0.73 |
| Time x drug | 0.001 | -0.025, 0.027 | 0.94 |
| Intervention x drug^1^ | 0.368 | -0.489, 1.225 | 0.40 |
| Intervention x drug x time^2^ | -0.058 | -0.241, 0.125 | 0.53 |
| **Number of prescriptions per 1000 population** |  |  |  |
| Intervention (post vs. pre) | -0.070 | -0.124, -0.017 | 0.01 |
| Drug (tirzepatide vs. semaglutide) | -0.005 | -0.057, 0.048 | 0.86 |
| Time | 0.045 | 0.041, 0.049 | <0.001 |
| Time x intervention | -0.013 | -0.020, -0.006 | 0.001 |
| Time x drug | -0.001 | -0.006, 0.003 | 0.59 |
| Intervention x drug^1^ | 0.020 | -0.057, 0.097 | 0.62 |
| Intervention x drug x time^2^ | -0.011 | -0.024, 0.002 | 0.11 |

1. Step change estimate: immediate change in prescribing rate at the time of the intervention for tirzepatide relative the same change for semaglutide

2. Slope change estimate: longer term change in trend in prescribing rates following the intervention for tirzepatide relative to semaglutide.

**Figure S3.** Controlled interrupted time series results for the rate of tirzepatide prescribing (number of prescriptions per 1000 population) compared to a control series (time-aligned, scaled semaglutide rates), based on English Prescribing Data for all of England combined

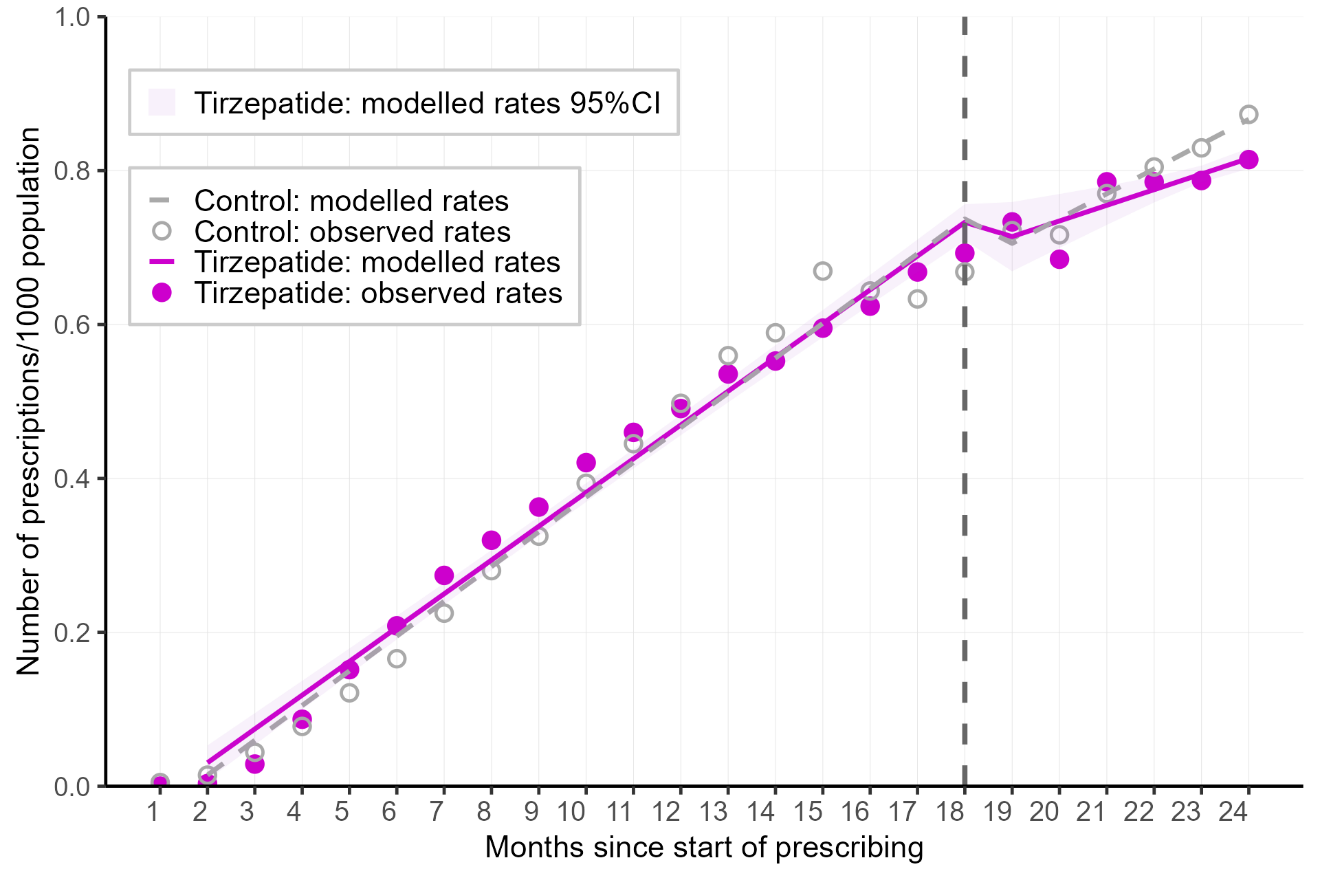

Note: Start of prescribing for tirzepatide in primary care was January 2024, and for semaglutide, January 2019.

**Figure S4.** Controlled interrupted time series point estimates with 95% confidence intervals assessing immediate step change and longer-term slope change over and above background trends, stratified by ICB. The step change estimates represent the estimated change in tirzepatide prescribing rates immediately following the intervention in June 2025, relative to changes in semaglutide prescribing rates. The slope change estimates represent the change in trend for tirzepatide prescribing rates in pre- versus post-intervention periods, relative to the trend change for semaglutide rates.

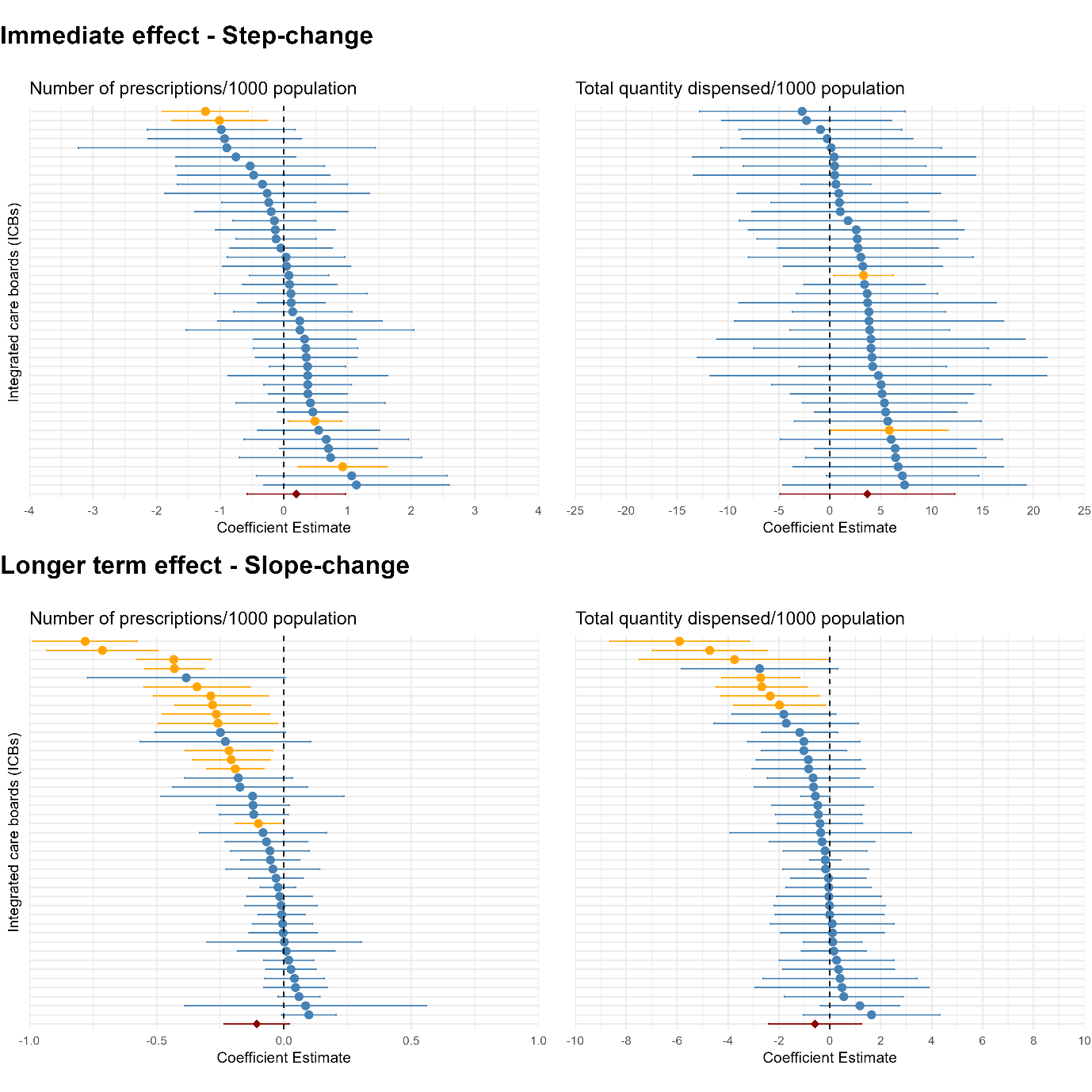
